# An optimal network that promotes the spread of an advantageous variant in an SIR epidemic

**DOI:** 10.1101/2024.10.25.24316154

**Authors:** Samuel Lopez, Natalia L. Komarova

## Abstract

In the course of epidemics, the pathogen may mutate to acquire a higher fitness. At the same time, such a mutant is automatically at a disadvantage because the resident virus has a head start in accessing the pool of susceptible individuals. We considered a class of tunable small-world networks, where a parameter, *p* (the rewiring probability), characterizes the prevalence of non-local connections, and we asked, whether the underlying network can influence the fate of a mutant virus. Under an SIR model, we considered two measures of mutant success: the expected height of the peak of mutant infected individuals, and the total number of recovered from mutant individuals at the end of the epidemic. Using these measures, we have found the existence of an optimal (for an advantageous mutant virus) rewiring probability that promotes a larger infected maximum and a larger total recovered population corresponding to the advantageous pathogen strain. This optimal rewiring probability decreases as mean degree and the infectivity of the wild type are increased, and it increases with the mutant advantage. The non-monotonic behavior of the advantageous mutant as a function of rewiring probability may shed light into some of the complex patterns in the size of mutant peaks experienced by different countries during the COVID19 pandemic.

## 1 Introduction

The study of the spread of infectious diseases is of great importance to ensure the health and security of a population. Susceptible-infected-recovered (SIR) models are a popular tool to study infections. Interesting insights can be gained by considering SIR dynamics in structured populations. By considering the spread of infectious diseases on a network with different tuneable features such as average node degree and cliquishness, one can better understand the effect that the host structure has on the spread of infectious diseases [48, 27].

In this study we will focus on network models of populations of hosts, and consider evolutionary aspects of infection propagation. Many disease-causing pathogens have been reported to mutate to obtain more efficient human-to-human transmissibility [34, 45]. Throughout the course of an epidemic, novel mutations may occur that change a fundamental characteristic of the infection; it may become more infectious, be infectious longer, or more fatal to name a few [31, 7]. For example, in 1918-1919 the world experienced a change in the fatality of the disease, with a mutation in H1N1 virus (the Spanish Flu). The wild type that led to the first wave had a fatality rate of 0.21% while a mutant strain leading to a second wave had a fatality rate of 2 − 4% [7]. The Zika virus (ZIKV) has been known to infect humans since the 1940’s. A mutation in 2013 likely contributed to a higher infectivity and microcephaly rate in humans [35, 54]. In 2013 a mutation of Ebola virus (EBOV), likely increasing the infectivity from the wild-type, contributed to the largest outbreak in West Africa [19, 50]. Further well known examples of mutations come from the SARS-CoV-2 (COVID19), which was first discovered in late 2019 [51]. By March 2023 there were more than 600 million total cases and 6 million deaths [1]. As the virus evolved, different strains became dominant. Notably widespread variants were the delta variant, which was the dominant strain in the summer of 2021, and the omicron variant which was the dominant strain in early 2022 [9, 4]. These strains were characterized by an increased infecivity [9].

A number of models of pathogens with mutant variants have been studied. Empirically, in [9], the takeover of a successful mutant in an SIR type model was considered, and it was shown that the turnover of the COVID-19 variants behaved similar to genetic sweeps in ecology, satisfying a logistic-type equation. In this paper, the dynamics of infection spread was described by using ordinary differential equations. In [13] a SIR type model with mutations was studied showing the large influence infectivity duration plays in replacement probability of an invading mutant. In [41] a two strain model with one strain following SIS dynamics and the mutant strain following SIR dynamics was shown to have asymptotically stable disease free equilibrium. Paper [14] formulated and studied and optimal control problem to find optimal treatment and quarantine strategies for a two strain infection. By coupling the SIR model with replicator dynamics they showed the optimal control could have a threshold structure. In paper [15] an SIR ODE model which incorporated vaccination and evolutionary game theory was studied. It was found that an early vaccination campaign is beneficial to a resident wild-type strain. Paper [23] modeled COVID19 with consideration of multiple vaccinations and mutant strains, highlighting the importance of vaccinating early to reduce the number of infected individuals. For other modeling approaches, see e.g. [16, 43, 18, 8, 29, 33, 42].

Dynamical modeling on networks has been also used to describe the spread of epidemics, including evolutionary aspects, where the host population structure is made more explicit [37, 44, 28, 12, 3, 25, 52, 55, 17, 24, 32, 53, 47]. Paper [27] studied a theoretical SIS infection with a mutant strain on empirical and theoretical networks. It was shown that network properties that promote the emergence of an original strain actually inhibit an invading mutant, and as a consequence the mutant displaces the wild-type less frequently. Paper [21] studied the influence the contact network has on the distribution of the number of “descendants” downstream from the initial “patient zero” mutant. Models of actively mutating pathogens spreading on networks have also been studied. Paper [39] investigated mutations that enhance the infection rate and found that the small-world property of the connection network could make the population more vulnerable to epidemic outbreaks, unless the number of long-range connections exceeded a threshold, reversing the effect. Paper [30] studied the spread of two competing viruses in a network by using an SIS infection model, and showed how mutations can affect the spreading ability of the virus and the coexistence of pathogens. Paper [40] considered an SIRS model on small world networks, demonstrating the existence of an interval of infectivity for the infection to achieve a non-extinct equilibrium. Pair-wise approximations are also a popular way to study a host-population structure, allowing for an ODE description of network effects [22, 26, 38, 20].

Infection like models on a network structure can also be extended to model non-biological contagion such as the spread of ideas (misinformation/rumors) and the spread of computer viruses. In [10], different network topologies were studied to determine their impact on the speed of the spread of computer viruses and worms. Paper [2] studied an SIRS model, which incorporated high and low influence layers, to describe information spread. It was shown analytically and through simulation that two information interference methods, targeted blocking and information dredging, are both effective ways to limit opinion spread. Other papers using SIR-like models to study the spread of non-biological contagion include [36, 46, 6].

In this paper we focus on the competition dynamics of two pathogen strains. We consider the spread of an infection on a random, static small-world network, and use the SIR model to study the co-dynamics of the wild-type and a mutant variant. To systematically vary the properties of the network, we adopt the framework of [27]. There, an important parameter is *p*, the rewiring probability, which controls the mean-shortest path length and the clustering coefficient of the network. Low values of *p* correspond to a lattice network with a significant neighborhood structure. A higher rewiring probability leads to the creation of non-local connections, with very high *p* values leading to the destruction of local neighborhoods and a global connectivity of the network. We ask whether such changes in the network structure may create conditions that are more (or less) favorable for spread of the mutant variant.

To this end, we characterize the effect that the network structure plays in the competition between a resident infection and an advantageous mutant variant. In particular, we focus on two measures of mutant efficiency: (i) ℐ_*m*_, the mutant peak infection (that is, the maximum number of individuals that are simultaneously infected by the mutant variant) and (ii) ℛ_*m*_, the total number of individuals that are recovered from the mutant variant by the end of the epidemic. We show that both ℐ_*m*_ and ℛ_*m*_ experience a peak as functions of *p*, the rewiring probability. In other words, there exist optimal networks that promote the largest infectious population and the largest recovered population by the mutant strain, which are between a regular ring lattice with a high clustering coefficient and large average distance between nodes, and a random network with no clustering and low average distance between nodes.

## 2 Results

### 2.1 The SIR model for two pathogen strains

In the standard SIR model, the population of hosts is categorized into three groups, the susceptible population *S*, the infected population *I*, and the removed population *R*. To consider the case when there are two strains of infection, a wild type and a mutant, the population is divided into 5 categories: the susceptible population *S*, the population infected with the wild-type *I*_*w*_, the population infected with the mutant *I*_*m*_, the population recovered from the wild-type *R*_*w*_, and the population recovered from the mutant *R*_*m*_. The rates for infection and recovery may be different for the two strains [31] and are given by parameters *β*_*w*_,*γ*_*w*_ and *β*_*m*_, *γ*_*m*_, which represent the infection and recovery rates for the wildtype and mutant respectively. Assuming there is no co-infection and recovery from either strain provides permanent immunity to both strains of infection, individuals progress through the categories as follows:

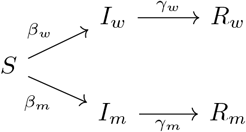

The spread of infection for the wild type and mutant strains can be modeled by the following system of ODE’s:

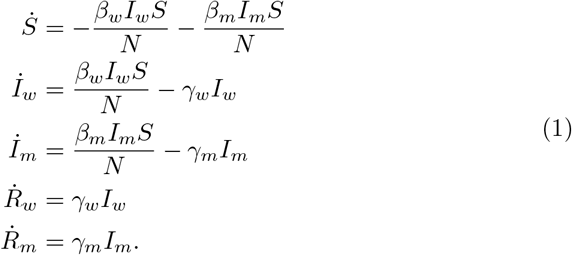

We create a stochastic model of an infection spreading through a network in the following way. For a given, fixed network with individuals as nodes, we assume that the nodes could be of five types corresponding to the five states introduced above. We further fix the per-time step probabilities of the different processes, *β*_*w*_, *β*_*m*_, *γ*_*w*_, *γ*_*w*_ to be constants between zero and one. We start by assuming that all the nodes are susceptible, and then randomly select a node to introduce the wild-type infection. For each time step, the network is randomly sampled until *M* infectious nodes have been sampled where *M* is the number of infectious nodes at the start of the time step. When an infectious node is sampled it has an opportunity to infect susceptible neighboring nodes, and does so with probability *β*_*w*_ per susceptible neighbor, then the node recovers with probability *γ*_*w*_. It is possible to select the same node more than once assuming the node did not recover in the prior selections.

If a sufficient portion (usually, .5% of the total population) of the nodes is simultaneously infected, then a mutant is introduced via one of the newly infectious nodes. This process of initialization represents a spontaneous mutation that occurs in the spreading pathogen. When there are two strains on the network, the algorithm is similar. The network is sampled *M*_*w*_ + *M*_*m*_ times, where *M*_*w*_ and *M*_*m*_ are the number of nodes infected by the wild-type and mutant respectively at the start of the time step. In this study we assume that the mutant is always advantageous, by setting *β*_*m*_ *> β*_*w*_ and *γ*_*m*_ = *γ*_*w*_. Nodes will only infect other nodes with the strain they carry, that is, no further mutations are considered. There is no co-infection, and recovery from either strain provides permanent immunity to both strains. The model runs until both strains are extinct.

Simulations that fail to introduce the mutant, that is, those runs where the number of infected individuals never rises above the threshold percentage of the total population, are discarded. Simulations where the mutant fails to spread (maximum of infectious with mutant *<*10) are also discarded. Simulations are aligned at the time of mutant introduction. Many independent runs are performed to obtain the means and standard deviations of the populations.

Here we focus on two quantities that characterize the mutant spread:

i. The mutant peak infection, defined as

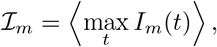

where the angular brackets denote the ensemble average.
ii. The total number of individuals that are recovered from the mutant variant by the end of the epidemic, defined as

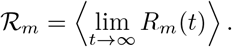

In this study, we explore a class of “small world” networks. Small world networks were first introduced by Watts and Strogatz as a way to create models between the two network extremes of a regular ring lattice and a random graph [49]. These networks are constructed by starting with a ring of equally spaced nodes and connecting them to their *k* nearest neighbors. Then edges are rewired with a tuneable rewiring probability *p*, creating “highways” between otherwise distant nodes. We define the distance between two nodes as the minimal number of edges between them [49]. Figure 1 shows three examples of small world networks with different values of the rewiring probability, *p*. In panel (a), *p* = 10^−4^ and we see a few “highways” allowing for large jumps in distance but the network has a large average distance (≈225). Then for *p* = 10^−2.4^ (panel (b)) we see much more of these “highways” and a much lower average distance between nodes (≈15). Finally, in panel (c) we have *p* = 1, which corresponds to an even lower average distance (≈3.7).

**Figure 1:**
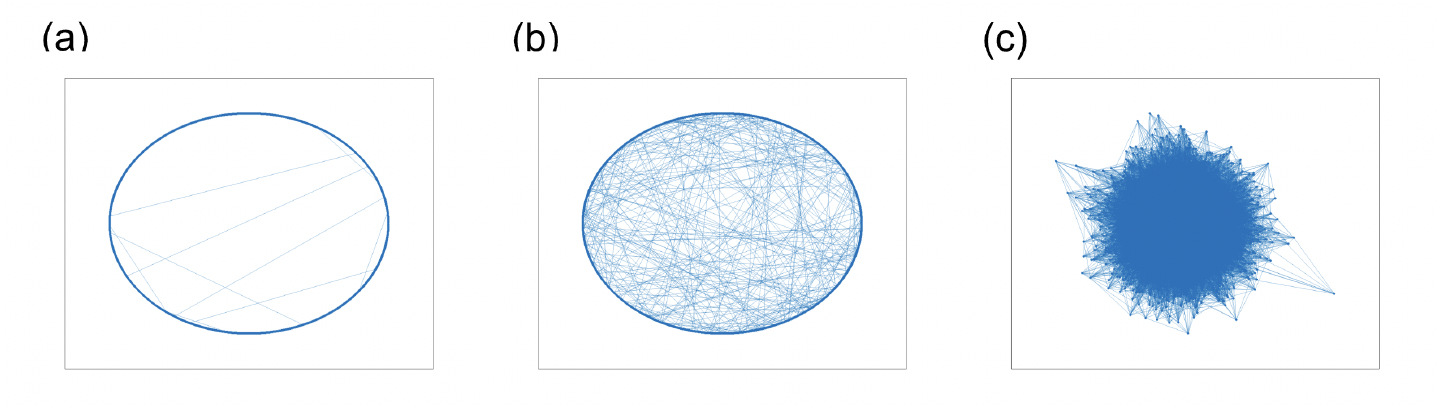
Typical graphs for *N* = 10, 000 and ⟨*k*⟩ = 14 for different rewiring probabilities, *p*. (a) *p* = 10^−4^, (b) *p* = 10^−2.4^, (c) *p* = 1.

Note that all the networks constructed by changing the parameter *p* are characterized by the same number of edges, or the mean degree.

### 2.2 The non-monotonic dependence of the peaks of mutant infection

We explore the infection trajectories for the individuals infected with wild type and mutant virus, for a range of rewiring probabilities, *p* = 10^*r*^ for *r* ∈ [− 2.8, 0] with 0.4 step sizes. Using the exponential scale for *p* allows us to better see the change in infection spread as more “highways” are added to the network. Figure 2 shows the mean trajectories of the populations infected with the wildtype and mutant, along with the standard error, which is shown by the shading. For each parameter combination, the number of infected individuals first rises as a function of time, then reaches a peak and declines toward zero, in accordance with the usual SIR epidemic dynamics.

**Figure 2:**
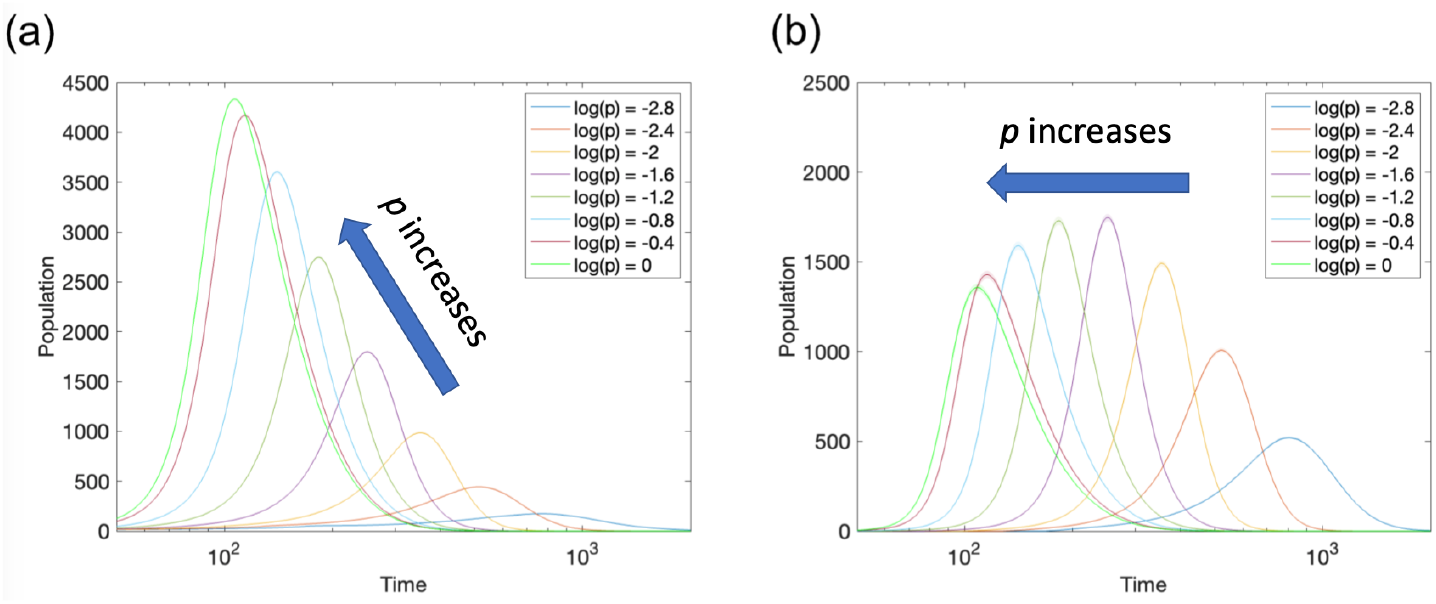
The mean, over *n* = 1999 simulations, of infectious population as a function of time for the wild type and mutant in panels (a) and (b) respectively. The shading (hard to see) represents the standard error. Different rewiring probabilities are shown by different colors according to the legend. The fixed parameters are *N* = 10, 000, ⟨*k*⟩ = 14, *β*_*w*_ = .01, *β*_*m*_ = .015, *γ* = .02.

Panel (a) of figure 2 shows the number of individuals infected with the wild-type; the different curves correspond to different values of the rewiring probability. Again, we note that for all the values of *p*, the mean degree of the network is preserved, and the changes affect the balance between local and global network structure. We observe a consistent increase in the height of the peak of infection as rewiring probability increases, along with a shift to the left, corresponding to earlier and earlier infection peaks. In other words, as more long-haul connections are incorporated in the network, the infection spreads faster and reaches a higher peak, which is an intuitive result.

Panel (b) of figure 2 gives the trajectories of the population infected with the mutant. As with the peaks of the wild-type infection, the mutant peak locations shift toward earlier times in the course of the epidemic, as *p* increases. Surprisingly, the peak height of mutant infection is non-monotone. The peak of mutant infection first increases as rewiring probability increases for lower *p* values. However, after the rewiring probability increases beyond a certain value, the peak starts decreasing, while shifting to the left all along. This suggests that there exists an intermediate, optimal value of the rewiring probability that facilitates the spread of advantageous mutants that are generated. This rewiring probability corresponds to an optimal network for an invading mutant strain to achieve a higher peak of infection. Simulation results for other parameter values, as well as information on the variance in the values, are presented in Appendix A.

### 2.3 Parameter dependence of the mutant peaks

Let the mean of the infection maximum be given by

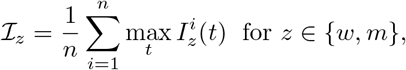

where the superscript enumerates independent simulation runs, and *n* is the number of simulations. Figure 3 shows ℐ_*m*_ as a function of rewiring probability with standard error bars, for a range of parameters. We explore how changes in network and infection parameters change the peak of maximum infectious with the mutant. An immediate effect of increasing the mean degree of the network is more “highways” around the network at lower rewiring probabilities. Figure 3(a) shows the mean of infectious maxima for different mean degrees: blue for ⟨*k*⟩ = 14, orange for ⟨*k*⟩ = 20, and yellow for ⟨*k*⟩ = 100. There is a clear shift in peak to a lower rewiring probability as the mean degree increases. Figure 3(b) fixes the mean degree ⟨*k*⟩ = 14 and varies the infectivity of the wild-type, *β*_*w*_, and/or the degree of mutant advantage, 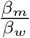. The blue line corresponds to *β*_*w*_ = .01 and *β*_*m*_ = 1.5*β*_*w*_. The orange line shows results for a system with an increased wild-type infectivity, *β*_*w*_ = .02 (under the same mutant advantage, *β*_*m*_ = 1.5*β*_*w*_); there is a shift in the mutant peak to a lower rewiring probability when compared to the blue line, which bears a resemblance to the case ⟨*k*⟩ = 20, *β*_*w*_ = .01 and *β*_*m*_ = 1.5*β*_*w*_ in panel (a). The purple line corresponds to the same wild-type infectivity as the blue line (*β*_*w*_ = .01), but the mutant infectivity is increased: *β*_*m*_ = 2*β*_*w*_. The mean of infection maximum for the mutant virus is now shifted to the higher rewiring probabilities, and it is also much shallower; this suggests the existence of a minimum mutant advantage 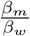 so a plateau is observed rather than a peak. Finally, the yellow line shows the results for a system where both wild-type and mutant infectivity has been increased: *β*_*w*_ = .02 and *β*_*m*_ = 2*β*_*w*_. We observe a shallower peak at the same location as the blue line.

**Figure 3:**
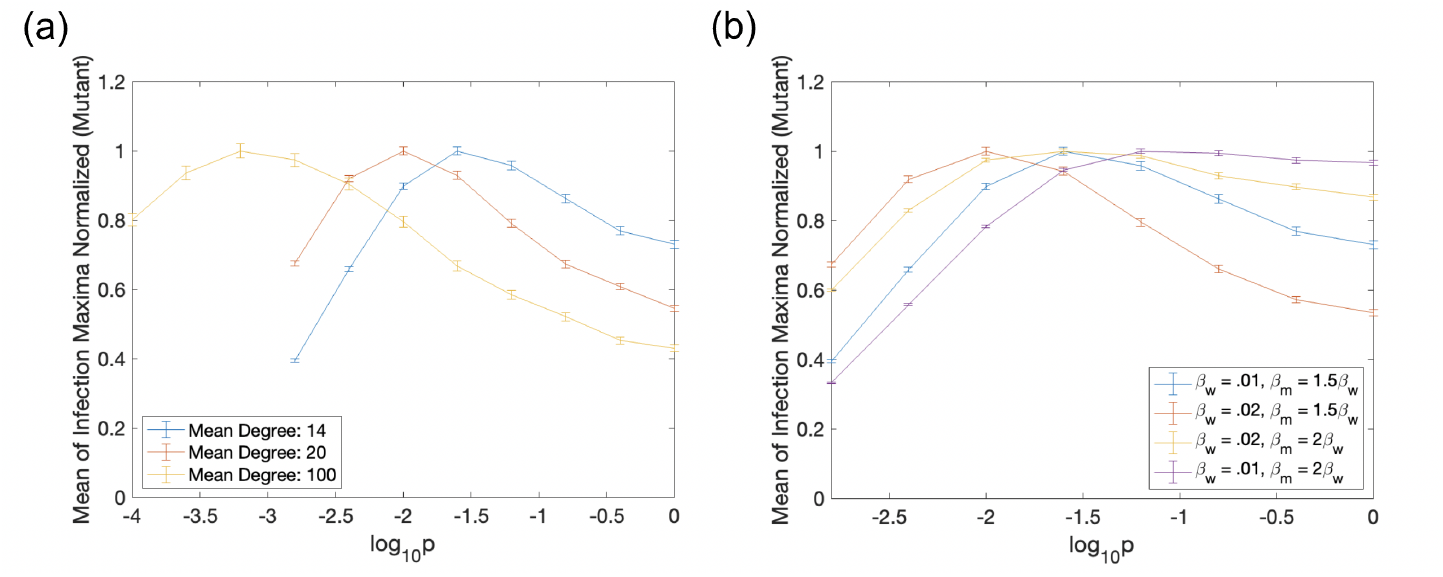
The mean, over *n* = 1999 simulations, of infectious maxima normalized as a function of rewiring probability for different parameter triples (⟨*k*⟩, *β*_*w*_, *β*_*m*_) with standard error bars. In panel (a) mean degree is varied according to the legend. The fixed parameters are *N* = 10000, *β*_*w*_ = .01, *β*_*m*_ = .015, *γ* = .02. In panel **b** infection rates *β*_*w*_ and advantage 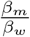 according to the legend. The fixed parameters are *N* = 10, 000, ⟨*k*⟩ = 14, *γ* = .02

Here we explore the dependence of the mutant peak on three parameters, the mean degree of the network, ⟨*k*⟩, the infectivity of the wild-type, *β*_*w*_, and the infectivity of the mutant, *β*_*m*_. Denote the location of the mutant peak for a given triple (⟨*k*⟩, *β*_*w*_, *β*_*m*_) as

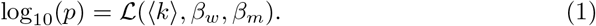

The above simulations show that ℒ(⟨*k*⟩, *β*_*w*_, *β*_*m*_) decreases with ⟨*k*⟩ and *β*_*w*_, and increases with *β*_*m*_.

To perform a more systematic study of this phenomenon, we have run extensive simulations varying the three parameters, and determined the location of the peak, ℒ (⟨*k*⟩, *β*_*w*_, *β*_*m*_). The peak of infectious maxima for the various parameter sets is found using the following algorithm. First, the mean of infectious maxima for a starting rewiring probability 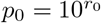 is computed. Then a step size is applied to the rewiring probability giving 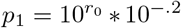 and the mean of infectious maxima is computed at the rewiring probability *p*_1_. If the mean of infection maxima at *p*_1_ is larger than the mean of infection maxima at *p*_0_ another step is taken *p*_2_ = *p*_1_ ∗ 10^−.2^. This is repeated until the mean of infection maxima at *p*_*i*+1_ is less than the mean of infection maxima at *p*_*i*_ which is the optimal rewiring probability for the corresponding parameter set.

To perform data fitting, we set

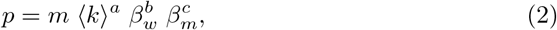

such that

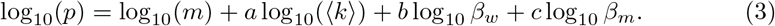

This allows us to perform linear regression of the data obtained from the described algorithm to find the coefficients *a, b*, and *c* in equation 3 and ultimately the exponents in equation 2. Using the method of least squares we find. The signs of the exponents are *a* < 0, *b* < 0, *c* > 0, which is consistent with what we observed in the graphed examples. The negative signs of *a* and *b* show the optimal rewiring probability for peak mean of infectious maxima occurs at a lower rewiring probability as (i) the network’s mean degree and (ii) wild-type infectivity *β*_*w*_ increase. The positive sign of *c* shows that the optimal rewiring probability increases as the mutant advantage increases.

Figure 4 plots the simulation results for a wide range of parameters together with the curve obtained from the best fit formula. The images illustrate the dependence of optimal rewiring probability for peak mean infectious maxima on the three parameters ⟨*k*⟩, *β*_*w*_ and *β*_*m*_. Specifically, for each plot we fix the mutant advantage, *β*_*m*_*/β*_*w*_, to be 1.25 in panel (a), 1.5 in panel (b) and 2.0 in panel (c), and plot the optimal rewiring probability, log_10_ *p* as a function of the network’s mean degree, for different values of the wild-type infectivity, *β*_*w*_.

**Figure 4:**
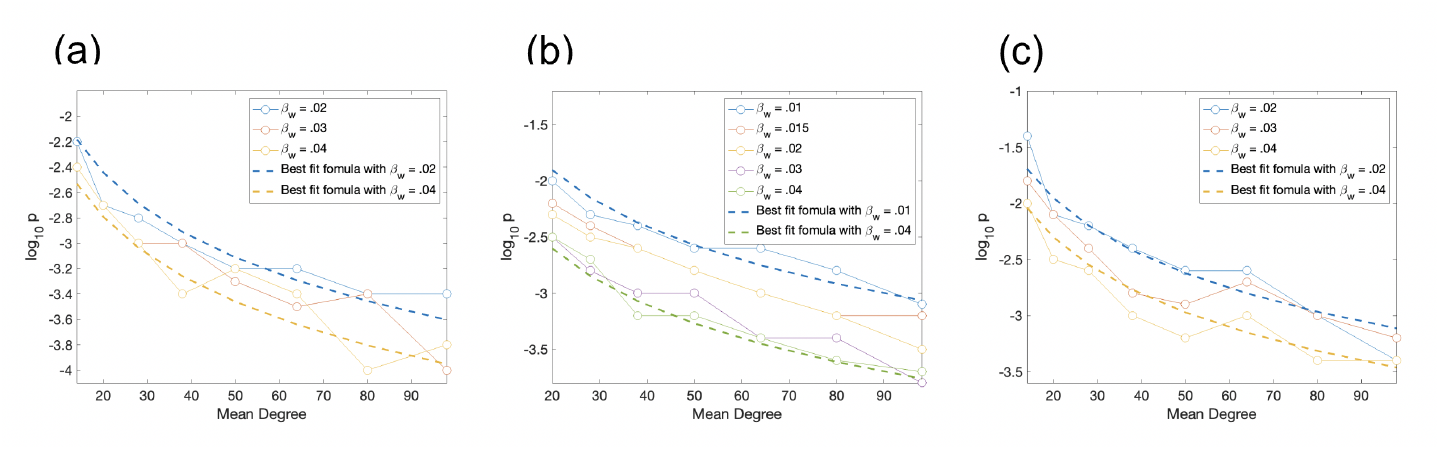
The optimal rewiring probability for largest peak of mutant infection. In all panels log_10_ *p* is plotted against mean degree, with *β*_*w*_ values following the legend. The data points for a parameter set are shown by the marks on the lines. Formula 2 with best fit parameters (log_10_(*m*) = −2.4557, *a* = −1.6816, *b* = −3.5538, and *c* = 2.3949) is shown by dashed lines. In panel (a) a low mutant advantage *β*_*m*_ = 1.25*β*_*w*_ taken, panel (b) shows a moderate mutant advantage *β*_*m*_ = 1.5*β*_*w*_, and panel (c) shows large mutant advantage *β*_*m*_ = 2*β*_*w*_.

### 2.4 The total number of recovered individuals

Next we consider the total number of nodes infected by either the wild type or the mutant by the end of the epidemic (that is, the number of individuals recovered from each infection type, as *t* → ∞). Let the mean of the recovered individuals be given by

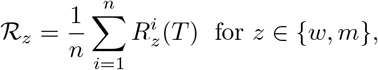

where the superscript enumerates independent simulation runs, *T* is the end of the simulation (when there are no more infected individuals), and *n* is the number of simulations.

Since we are considering SIR dynamics, there is no movement out of the recovered state (in contrast to the set of infectious nodes, which gains members as susceptible nodes become infected but also loses members as nodes recover). Figure 5(a) shows the average number of nodes that recovered from being infected by the wild-type, as a function of rewiring probability. The curves tend to be increasing as rewiring probability increases, with the exception of some parameter sets at very low rewiring probability; for those parameter sets, the curves reach a minimum value for relatively low *p* values.

**Figure 5:**
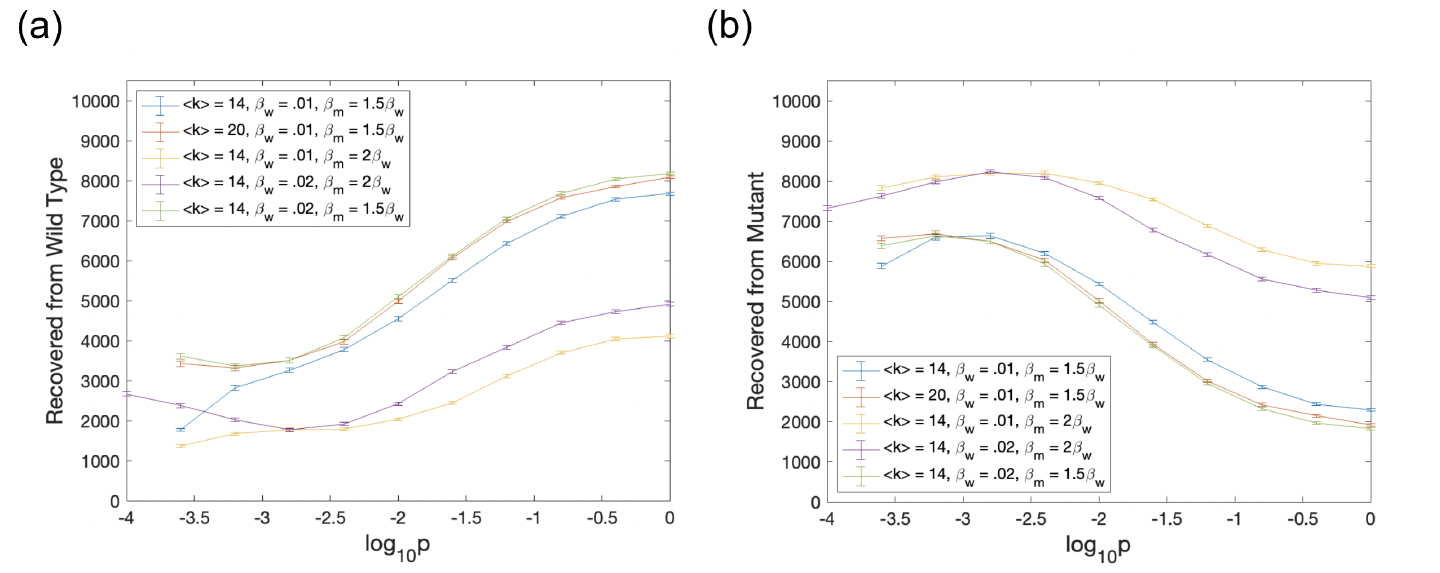
The mean, over *n* = 1999 simulations, of number of nodes recovered from the wild type in panel (a) and recovered from the mutant in panel (b) with standard error bars. Parameter triples (⟨*k*⟩, *β*_*w*_, *β*_*m*_) are varied according to the legend. The fixed parameters are *N* = 10000 and *γ* = .02

Figure 5(b) shows the average number of nodes that recovered from being infected by the mutant as a function of rewiring probability. As with the infection peaks, the number of recovered individuals exhibits a qualitatively different behavior for the mutant strain, compared to the wild-type infection. All the curves for mutant strain appear non-monotone, achieving a maximum at a relatively low rewiring probability.

We observe that the peak of infectious population vs the peak of total recovered do not coincide at the same rewiring probability. The peak for total recovered, ℛ_*m*_, typically corresponds to a lower rewiring probability, compared to ℐ_*m*_. Qualitatively, however, it has properties similar to the peak of mutant infection. Namely, the optimal value of the rewiring probability that maximizes R_*m*_ decreases with ⟨*k*⟩ and *β*_*w*_, and it increases with *β*_*m*_.

### 2.5 The optimal (for mutant) rewiring probability

The above observations suggest that in the range between a regular lattice (*p* = 0) and a random network without any neighborhood structure (*p* = 1), there is an optimal rewiring probability that maximizes the spread of an advantageous mutant. This optimal rewiring parameter is obtained as a trade-off of several factors that limit the spread of a mutant.

On the one hand, very low rewiring probabilities make the infection spread very ineffective, and thus increasing *p* from very low numbers will lead to an increase in mutant spread. On the other hand, very large values of rewiring probability are disadvantageous for the mutant, because they allow a very quick wild-type epidemic spread, such that the mutant does not get a chance to catch up, despite its increased infectivity. A trade-off of these two factors leads to the existence of an optimal network structure that favors mutant spread. Below we will analyze these two trends in detail.

#### Very low rewiring probabilities make the infection spread very ineffective

This is true for any infection, and for a mutant in particular. To see this, we note the following patterns of epidemic spread behavior at low values of *p*.

- At very low rewiring probability, the epidemic can be very “inefficient”, such that the total number of recovered individuals at the end is less than the number of nodes, *R*_*m*_ + *I*_*m*_ *< N*, see figure 6(a), where the number of nodes recovered from the mutant (blue), the number of nodes recovered from the wild type (orange) and the total number of nodes recovered (black) are plotted together for a single set of parameters. In this case, ℛ_*m*_ and ℛ_*w*_ increase with *p* initially.
- For other parameter combinations (particularly when *β*_*w*_ or mean degree is larger), it is possible that the total number of recovereds *R*_*m*_+*I*_*m*_ = *N* even for very low *p*, but an increase in the rewiring probability will still benefit the mutants (possibly at the expense of the wild types). This is illustrated in figure 6(b), which shows an increase in ℛ_*m*_ and a simultaneous decrease in ℛ_*w*_ as *p* first increases. At *p* = 0, the mutant may be stuck in one spatial location, as a consequence of its initial placement; the wild-type is more spread due to its head-start in the epidemic. As long-haul connections are added, this opens up new locations for the mutant, thus reducing this discrepancy.

**Figure 6:**
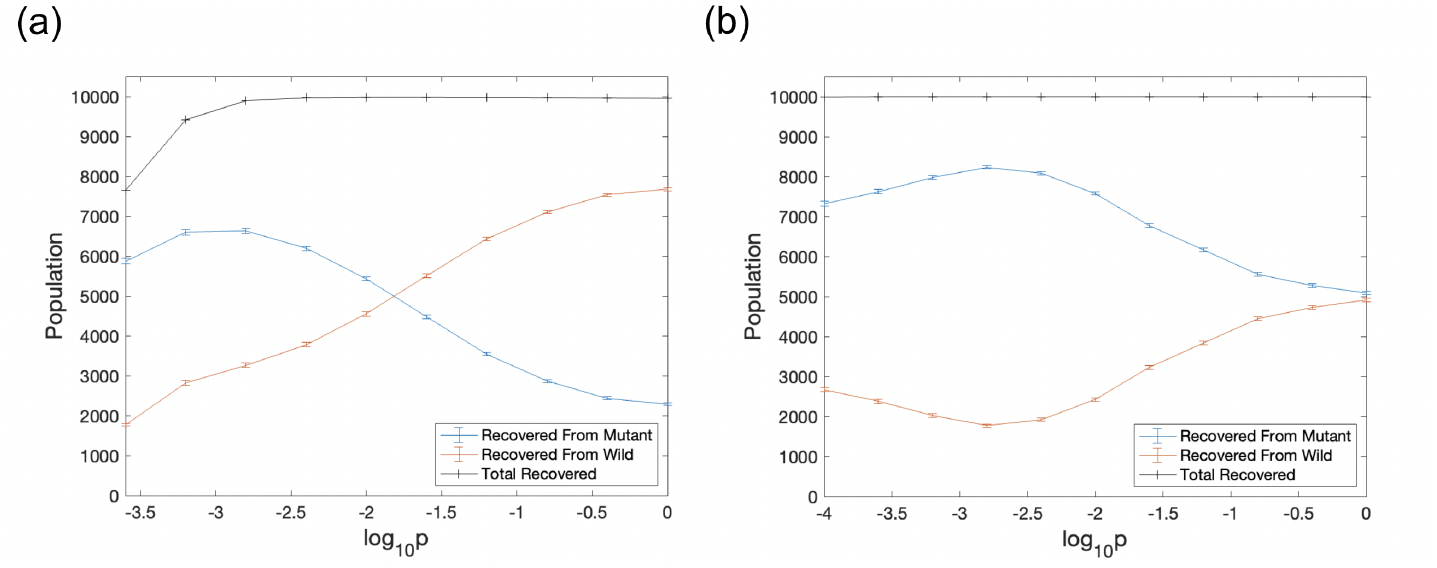
The mean, over *n* = 1999 simulations, of number of nodes recovered from the wild type, mutant and total infected nodes with standard error bars. In panel (a) the parameter triple is (⟨*k*⟩, *β*_*w*_, *β*_*m*_) = (14, .02, .04). In panel (b) the parameter triple is (⟨*k*⟩, *β*_*w*_, *β*_*m*_) = (14, .01, .015). The fixed parameters are *N* = 10000 and *γ* = .02

Therefore, it is intuitively clear that an increase in the number of non-local edges will lead to an increase in the number of nodes available for mutant infection.

#### Very large values of rewiring probability are disadvantageous for the mutant

There is a clear decrease from a peak in ℐ_*m*_ for most parameter sets and ℛ_*m*_ for all parameter sets. This can be attributed to a single factor: a sort of “first movers” advantage. In our model the wild type is given an opportunity to spread in the absence of the mutant strain until ≈.5% of available nodes are simultaneously infected (for the networks prtesented here, *N* = 10, 000 so there is approximately 50 wild type nodes at the time step the mutant is introduced). Consequentially, the wild type is spreading faster, in the sense of infecting more nodes per time step, than the more infectious mutant when *I*_*m*_ is small. A key characteristic in small-world networks is the rapid decrease in mean distance between nodes as rewiring probability increases, see figure 7(a), which leads to a faster overall epidemic. Therefore, the advantage in numbers that the wildtype has is more impactful as rewiring probability increases.

**Figure 7:**
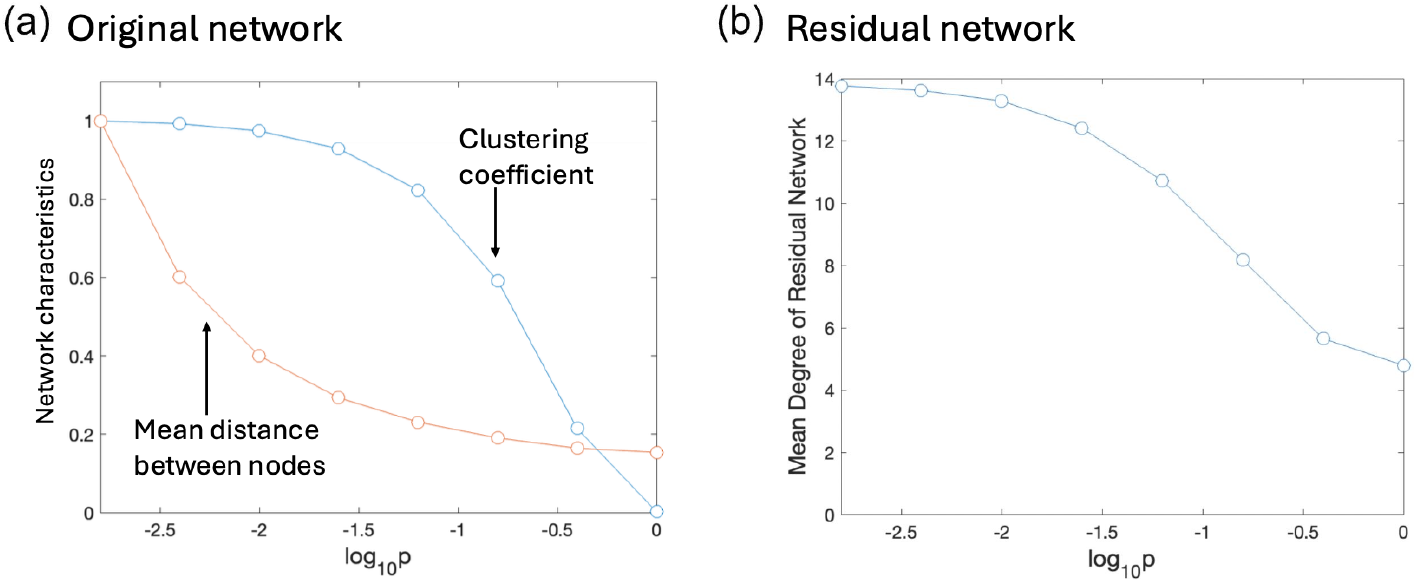
Network characteristics as a function of the rewiring probability, *p*: (a) the mean node distance and (b) the mean degree of the residual mutant network as functions of rewiring probability, with parameters *N* = 10000, ⟨*k*⟩ = 14, *β*_*w*_ = .01, *γ* = .02 and *β*_*m*_ = 1.5*β*_*w*_. The average distance between nodes quickly decreases while the mean degree of the residual mutant network begins very close to the mean degree of the original network and decreases as rewiring probability increases.

To see this it is insightful to study the residual mutant network, which is defined as the subnetwork obtained from the set of all nodes recovered from the mutant, at the end of the epidemic. Figure 7(b) shows the mean degree of the residual mutant network as a function of rewiring probability. At low rewiring probability the mean degree of the residual mutant network is very close to the mean degree of the original network of 14. This shows that at low rewiring probability, as the mutant infects a node, it also infects most of the neighbors of that node, moving very thoroughly throughout the network. As rewiring probability increases, the mean degree of the residual mutant network decreases. The mutant is less consistently able to infect a node and all of its neighbors.

There are both a “cost” and a “benefit” to increasing the rewiring probability, as the wild type and mutant are competing to infect susceptible nodes on a shared network. To see this, for each susceptible node with at least one mutant neighbor, *i*, denote by 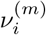 the number of edges that connect *i* with an infected mutant node, and by 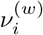 the number of edges that connect *i* with an infected wild-type node. Denote the ratio 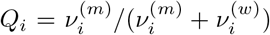, see an illustration in 8(a). There, susceptible nodes are blue, wild type nodes are red, and mutant nodes are black. There are three susceptible nodes connected to a mutant node. Two of these nodes’ infectious neighbors are exclusively mutant, yielding *Q*_*i*_ = 1. The remaining susceptible node with a mutant neighbor has one edge leading to a wild-type node and one edge leading to a mutant node. This node has *Q*_*i*_ = .5. Hence, for the schematic, the mean (over the three susceptible nodes) is 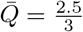. In addition to the quantity 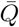, it is also useful to keep track of connections between wild-type infected and susceptible nodes (*S* − *I*_*w*_ Pair) and mutant infected and susceptible nodes (*S* − *I*_*m*_ Pair). In the example of figure 8(a), there are two wild type nodes and three edges, highlighted in red, connecting a susceptible nodes to a wild-type node; this would have a *S* − *I*_*w*_ Pair value of 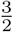. There is one mutant node and three edges, highlighted in black, connecting susceptible nodes to the mutant node; this would have a *S* − *I*_*m*_ Pair value of 3.

**Figure 8:**
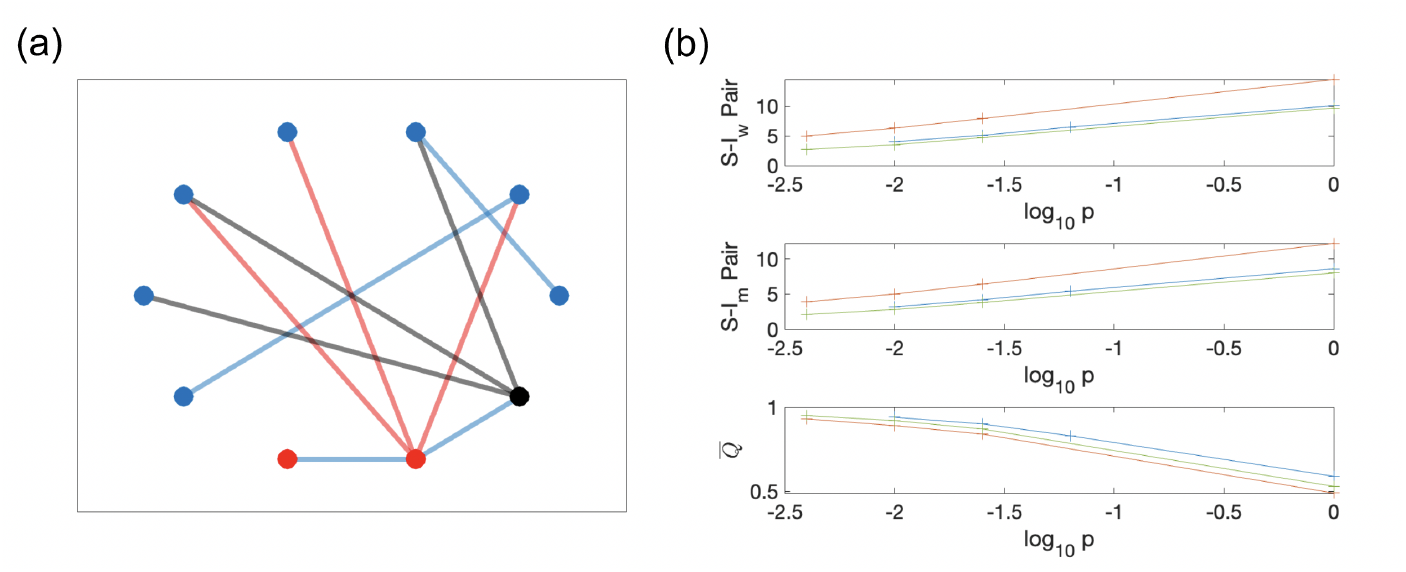
The costs and benefits of increasing *p*. (a) A schematic illustrating how the three measures, *S* − *I*_*w*_, *S* − *I*_*m*_ and 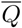, are calculated. Susceptible nodes are blue, wild type nodes are red, and mutant nodes are black. There are two wild type nodes and three edges, highlighted in red, connecting a susceptible nodes to a wild-type node; resulting in the *S* − *I*_*w*_ Pair value of 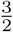. There is one mutant node and three edges, highlighted in black, connecting susceptible nodes to the mutant node; this means that the *S* − *I*_*m*_ Pair value is 3. There are three susceptible nodes connected to a mutant node. Two of these nodes’ infectious neighbors are exclusively mutant *Q*_*i*_ = 1. The remaining susceptible node with mutant neighbor has one edge leading to a wild-type node and one edge leading to a mutant node. This node has *Q*_*i*_ = .5. Hence, for the schematic 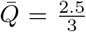. (b) The behavior of quantities *S* − *I*_*w*_ pair, *S* − *I*_*m*_ pair, and 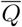 as functions of *p*, for parameter triples (⟨*k*⟩, *β*_*w*_, *β*_*m*_) given by (14, .01, .015) (blue), (20, .01, .015) (orange), (14, .02, .015) (green).

Figure 8(b) illustrates both costs and benefits of increasing the rewiring probability, from the mutant’s point of view. On the one hand, the number of *S* − *I*_*m*_ Pairs grows, as decreasing the average distance between nodes provides a steady supply of susceptible nodes for the mutant to infect. This can be seen in the middle graph (*S* − *I*_*m*_ Pair) of figure 8(b). On the other hand, the wild-type also gains the same “benefit” as the mutant (see figure 8(b) top graph, *S* − *I*_*w*_ Pair), while the mutant is losing a substantial local dominance, as seen in the bottom panel of Figure 8(b). There, we see the behavior of the quantity 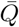 that is also time-averaged, as a function of the rewiring probability. The temporal average is found by first taking the average from the start of the epidemic to the peak of the mutant strain on a simulation by simulation basis, then averaging the simulation mean over the 999 simulations. We observe that this quantity decreases as *p* increases, suggesting that mutants start to “lose” the competition for the target susceptible individuals, over the wild-type infection.

To conclude, as the probability of rewiring, *p*, increases, the conditions for the spread of an advantageous mutant first improve and then become worse, leading to an intermediate regime that is the most favorable for mutant spread. This “sweet spot” fits in the window of rewiring probabilities where the clustering coefficient is still large, but the mean distance between nodes is already relatively small, see figure 7(a).

## 3 Discussion

When an advantageous mutant in generated by a spreading resident virus, it has both an advantage and a disadvantage compared to the resident virus population. The advantage is the higher infectivity (which we assume). The disadvantage is the fact that the resident virus has a head start, and the number of susceptible target individuals is shrinking. In this paper we asked, how do these two factors trade off, and whether the underlying network can influence the fate of a mutant virus.

We considered a class of tunable small-world networks, where a parameter, *p* (the rewiring probability), characterizes the prevalence of non-local connections, bridging between a perfect circle network at *p* = 0 and a completely random graph with no clustering or neighborhood structure, but a much reduced network diameter at *p* = 1. Under an SIR model, we considered two measures of mutant success: the expected height of the peak of mutant infected individuals (ℐ_*m*_), and the total number of recovered from mutant individuals at the end of the epidemic (ℛ_*m*_). Using these measures, we have found the existence of an optimal (for an advantageous mutant virus) rewiring probability that promotes a larger infected maximum and a larger total recovered population corresponding to an advantageous mutant. It was observed that the mutant will have a peak in (ℛ_*m*_) at a relatively low rewiring probability and a peak of (ℐ_*m*_) at a somewhat larger rewiring probability.

The reason for this intermediate optimal rewiring probability that favors mutants is as follows. As *p* increases, the virus gains easier and faster access to the whole network. Too small values of *p* prevent the mutant (which is initially in the minority) from spreading globally. Too high values of *p* result in the epidemic burning through the population too fast, such that the mutant cannot close the gap to the wild-type. One can think of this competition of the two viral strains as a race, with the mutant being a faster opponent but the wild type having a head start. A 90 meter head start in a 100 meter race is more influential than a 90 meter head start in a marathon.

The three parameters: mean degree, wild-type infectivity *β*_*w*_, and mutant advantage 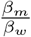 influence the value of optimum rewiring probabilities. Although the optimum rewiring probabilities for total recovered and peak of infectivity differed, they showed similar quantitative properties shifting to a lower rewiring probability as mean degree and *β*_*w*_ are increased and shifting to a higher rewiring probability as 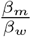 is increased. The directions of the shifts are intuitive when considering the causes of the peaks. As the mean degree increases, a lower rewiring probability is sufficient for the effective spread of the mutant to infect the whole network and to not leave the wild-type as the exclusive strain in a part of the network for many time-steps, leading to a decrease in optimal *p*. Increasing *β*_*w*_ also allows for the infection to spread to more nodes at lower *p*, though it likely has more influence on the ability to infect the whole network rather than the mutant’s ability to traverse the network to distant nodes. As mutant advantage 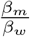 increases the mutant is more effective and requires less time to out-compete the wild-type, leading to an increase in optimal *p*.

Although our simulations pertain to a theoretical pathogen on theoretical networks, the logic of how a host populations structure influences the peak of infectious for an advantageous mutant may be extended to real epidemics. The world recently endured an epidemic caused by SARS-CoV-2 (COVID19), which contains a large number of variants of the virus. Here we focus on three variants: “alpha”, “delta”, and “omicron” because they were created consecutively. We will first treat the “alpha” variant as the wild-type and the “delta” variant as the advantageous mutant. We will then treat the “delta” variant as the wild-type and the “omicron” variant as the advantageous mutant. For illustration, we will consider three high-income Western countries: the US, Canada, and Denmark. These three SARS-CoV-2 variants were the dominant strains between April 2021 and April 2022 in the United States, Denmark, and Canada [1].

There are three distinct peaks in new cases corresponding to one of the three variants shown in figure 9 [1]. In panel 9(a), we see a peak in new cases caused by the delta variant with the United States having the largest peak and Denmark with a slightly larger peak than Canada. Interestingly, the peak during the omicron wave was largest for Denmark, then the US, and Canada with the smallest peak. However, omicron has a substantial peak in all countries compared to the delta variant, see panel 9(b).

**Figure 9:**
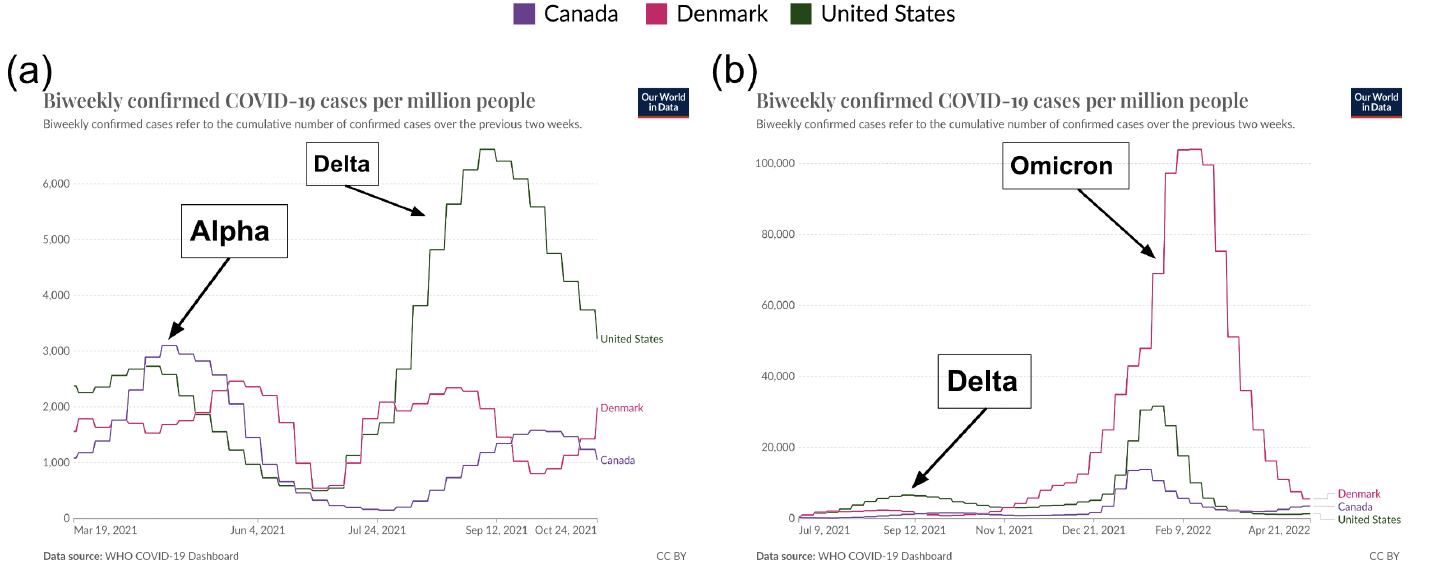
Biweekly confirmed COVID19 cases per million people in Canada, Denmark and United States with data from [1].

There are many factors to consider, to effectively translate into the rewiring probability, mean degree, wild-type infectivity *β*_*w*_, and mutant infectivity *β*_*m*_. Public policies such as the use of masks, social distancing, work-from-home, vaccination, and quarantine were some of the measures implemented by health officials and policymakers to limit the spread of COVID-19 during the pandemic. The various policies can reasonably be separated into either a real-world representation of “decreasing mean degree” or “decreasing infectivity.” Notably, we showed decreases to mean degree and infectivity (assuming it decreases infectivity of the wild-type and mutant with minimal impact on 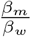) both increase optimal rewiring probability; thus, to illustrate our model, we do not need to classify each policy into one of the two categories. It is however important to consider the consequences of some of the restriction policies on the network structure. We claim that, in the context of our framework, restricting travel and other contacts served both to decrease the network’s mean degree, and to decrease its rewiring probability. Suppose a node has ten connections and 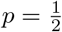, then we would expect five long-range connections. Next, consider policies that aim to decrease the mean degree particularly by removing these “long-range” connections. Our node now may be down to six (for argument’s sake) connections after efforts removed four long-range connections, leaving our node with five local connections and one long-range connection. For a network with mean degree 6, this is indicative of 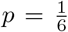, and not 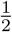. Therefore, we conclude that adoption of protective measures likely resulted in (1) a decrease in the mean degree, (2) a decrease in infectivity, and (3) a decrease in the rewiring probability.

The rise of the new SARS-CoV-2 variants was attributed to their increased infectivity. The “delta” variant was reported to be more infectious than “alpha” (between 1.6-2.6 times more transmissible), while the “omicron” variant was more infectious than the “delta” variant (approximately 3.3 times more transmissible) [11, 5]. In figure 10 we use the estimated values of the infectivity increase for the “delta” and “omicron” variants to plot the predicted optimal rewiring probability. The blue curve shows the optimal rewiring probability for “delta” and the green curve the optimal rewiring probability for “omicron”. The triangles represent a hypothetical scenario that could explain the differences in the peaks of the SARS-CoV-2 variants experienced by the three countries. This is more of a proof of principle rather than a parameter fitting exercise, which shows that our theoretical approach may elucidate some subtle reasons for observed virus behavior.

**Figure 10:**
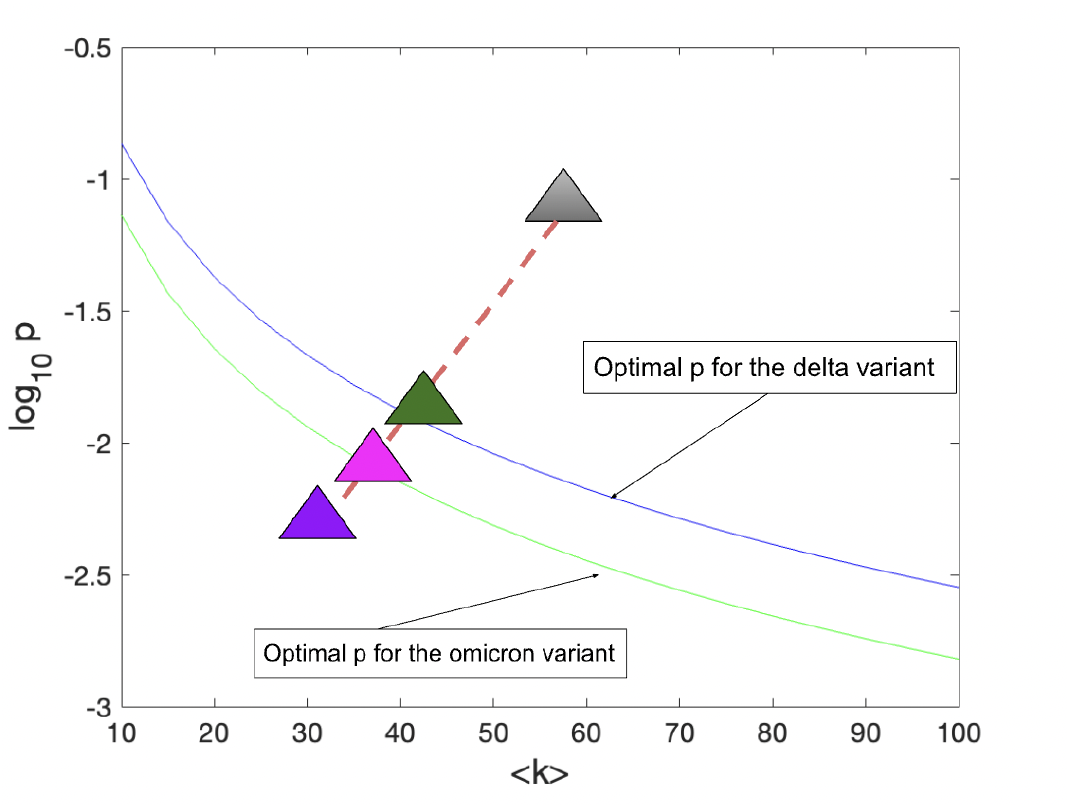
Two best fit curves from equation 3 with parameter values *β*_*w*_ = .01 and *β*_*m*_ = 2.5*β*_*w*_ in blue and *β*_*w*_ = .025 and *β*_*m*_ = 3*β*_*w*_ in green. The gray triangle is an arbitrary starting population structure when there are no public health policies and the three colored triangles represent where a population may land dependent on the adoption level of public health policies.

In the hypothetical scenario that we are considering, the gray triangle in figure 10 represents the pre-pandemic mean degree and the rewiring probability that characterized the contagion networks in all the three countries. Upon the start of the pandemic, all the countries implemented protective measures that led as we discussed, to a decrease in ⟨*k*⟩, infectivity, and the rewiring probability. This may have resulted in a trajectory shown by the red dashed line. The degree to which the three country’s populations complied with the restrictions may have been slightly different. In our example, the country represented by the purple triangle achieved the largest decrease in ⟨*k*⟩ and *p*, while the country represented by the green triangle had the smallest change. As a result, the “green” country (possibly illustrating the US in our example) may have moved to the position that is close to the optimal curve for the “delta” variant, and experienced the largest “delta” wave, while the “pinl” country (“Denmark”) shifted to the vicinity of the optimal curve for the “omicron”. The “purple” country” (representing Canada) is the furthest from the two optimal curves, which is consistent with its experiencing the mildest peaks of both variants.

Our methodology does not allow us to argue exactly what changes were experienced by different countries. Moreover, our model system (small-world networks, and the SIR model for the epidemic) is highly stylized. It is however instructive to study the behavior of the mutant in this simplified setting, because it gives rise to insights that can be applicable in more complex settings.

## Data Availability

All data produced in the present study are available upon reasonable request to the authors

## A Additional images of wild-type and mutant co-dynamics

In figure 2 we showed the mean over *n* = 1, 999 simulations of the infectious populations with standard error bars for one parameter set. Here, in figures 11-22, we show the mean over *n* = 1, 999 simulations for multiple different parameter sets including changes to mean degree ⟨*k*⟩ = 14, infectivity *β*_*w*_ and mutant advantage 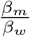 for the wild-type and mutant. Further, we show standard deviation as a shaded region in panel (a) while panel (b) shows standard error (generally hard to see).

**Figure 11:**
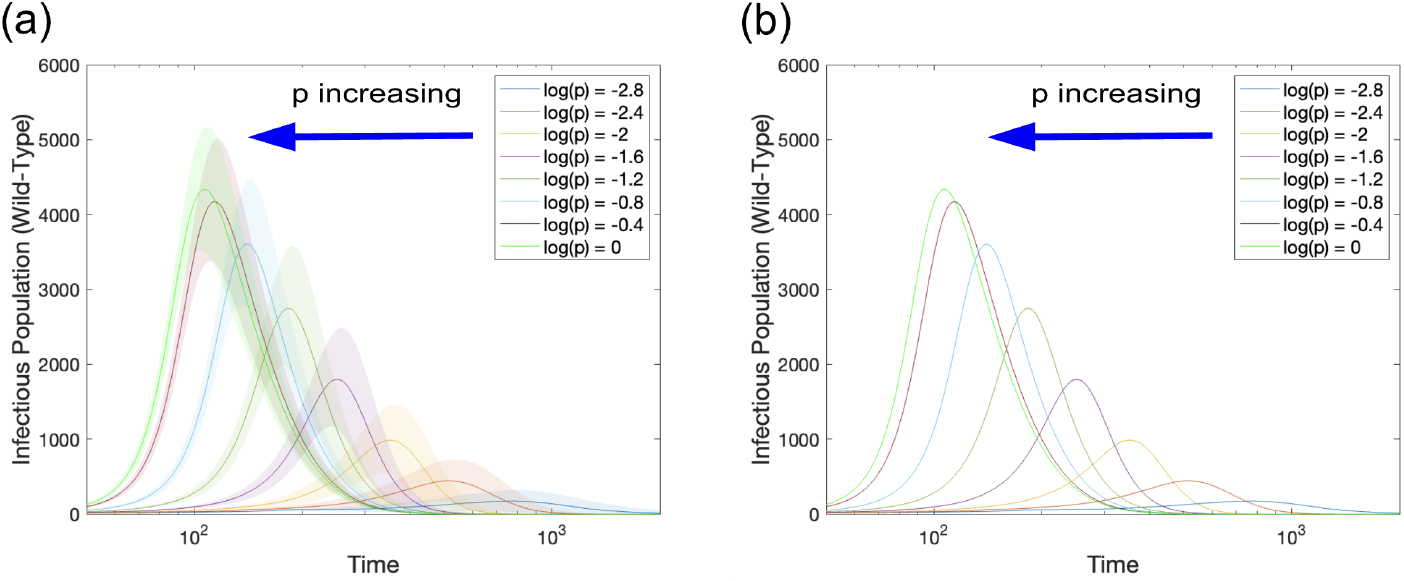
Mean wild-type infectious population for many *p* values. In both panels we show the mean wild-type infectious population over 1,999 simulations. In panel (a) the shaded region represents the standard deviation and in panel (b) the shaded region represents the standard error. Parameters are *N* = 10, 000, ⟨*k*⟩ = 14, *β*_*w*_ = .01, *β*_*m*_ = 1.5*β*_*w*_, *γ* = .02, and *p* is varied according to the legend.

Figures 11, 13 and 15 show the wild-type for mean degrees 14, 20 and 100 with other parameters equal. We see the wild-type virus producing a larger peak, which appears earlier as mean degree increases. However, there is more nuance in the mutant case shown in figures 12, 14 and 16. Take for example *p* = 10^−1.6^, then the mutant peaks at approximately 1800, 2100, and 1800 for mean degrees 14, 20 and 100 respectively, i.e. non-monotone.

**Figure 12:**
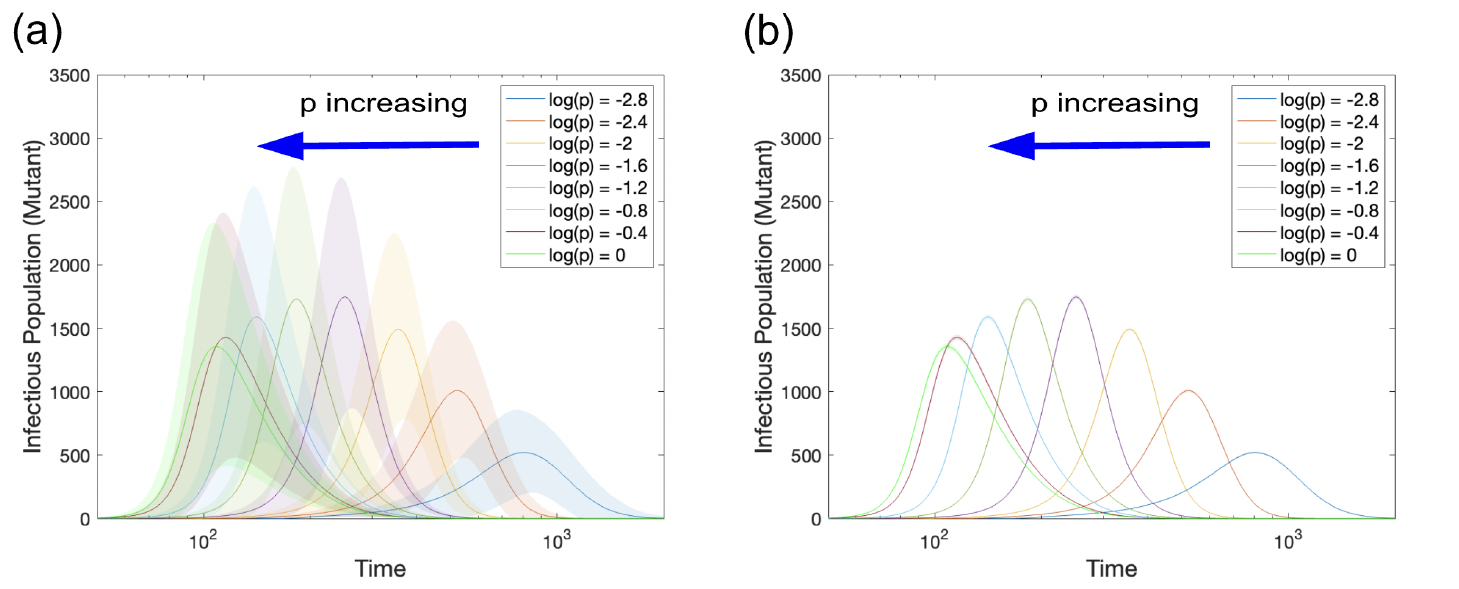
Mean mutant infectious population for many *p* values. In both panels we show the mean mutant infectious population over 1,999 simulations. In panel (a) the shaded region represents the standard deviation and in panel (b) the shaded region represents the standard error. Parameters are *N* = 10, 000, ⟨*k*⟩ = 14, *β*_*w*_ = .01, *β*_*m*_ = 1.5*β*_*w*_, *γ* = .02, and *p* is varied according to the legend.

**Figure 13:**
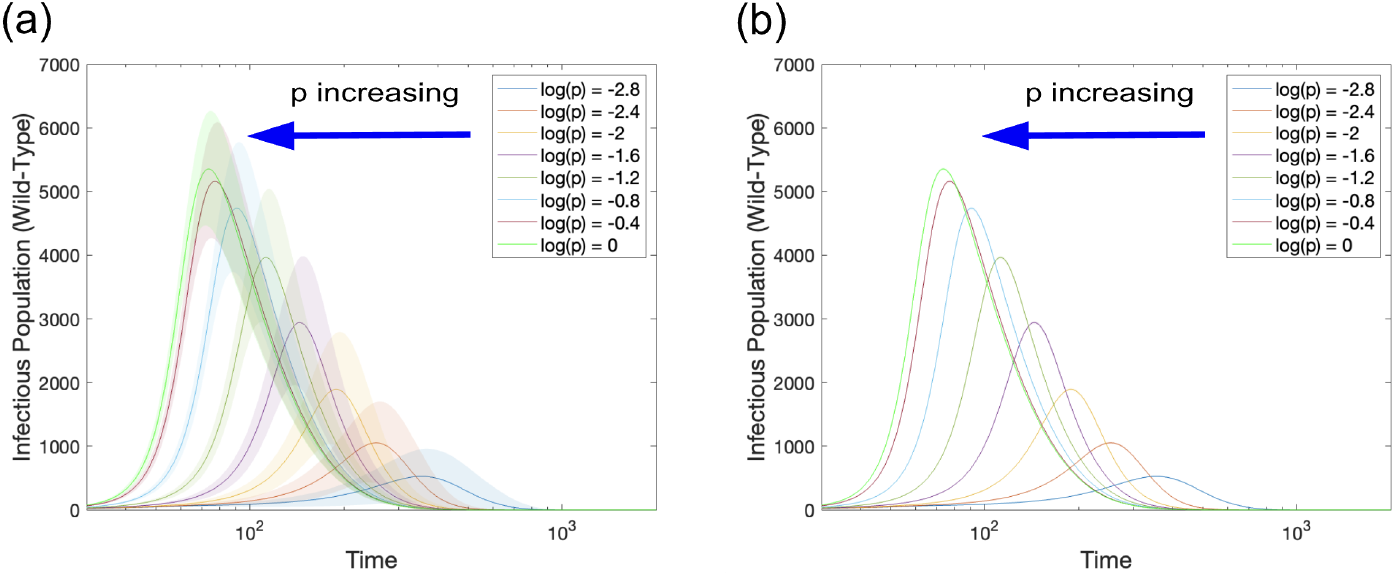
Mean wild-type infectious population for many *p* values. In both panels we show the mean wild-type infectious population over 1,999 simulations. In panel (a) the shaded region represents the standard deviation and in panel (b) the shaded region represents the standard error. Parameters are *N* = 10, 000, ⟨*k*⟩ = 20, *β*_*w*_ = .01, *β*_*m*_ = 1.5*β*_*w*_, *γ* = .02, and *p* is varied according to the legend.

**Figure 14:**
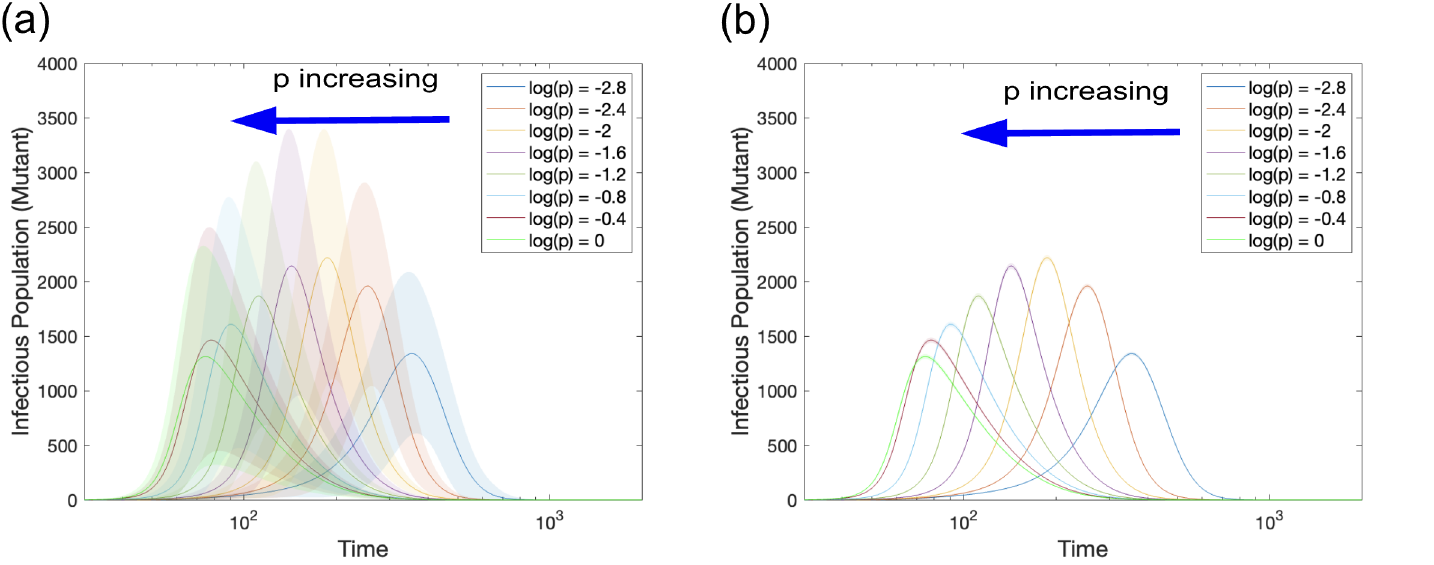
Mean mutant infectious population for many *p* values. In both panels we show the mean mutant infectious population over 1,999 simulations. In panel (a) the shaded region represents the standard deviation and in panel (b) the shaded region represents the standard error. Parameters are *N* = 10, 000, ⟨*k*⟩ = 20, *β*_*w*_ = .01, *β*_*m*_ = 1.5*β*_*w*_, *γ* = .02, and *p* is varied according to the legend.

**Figure 15:**
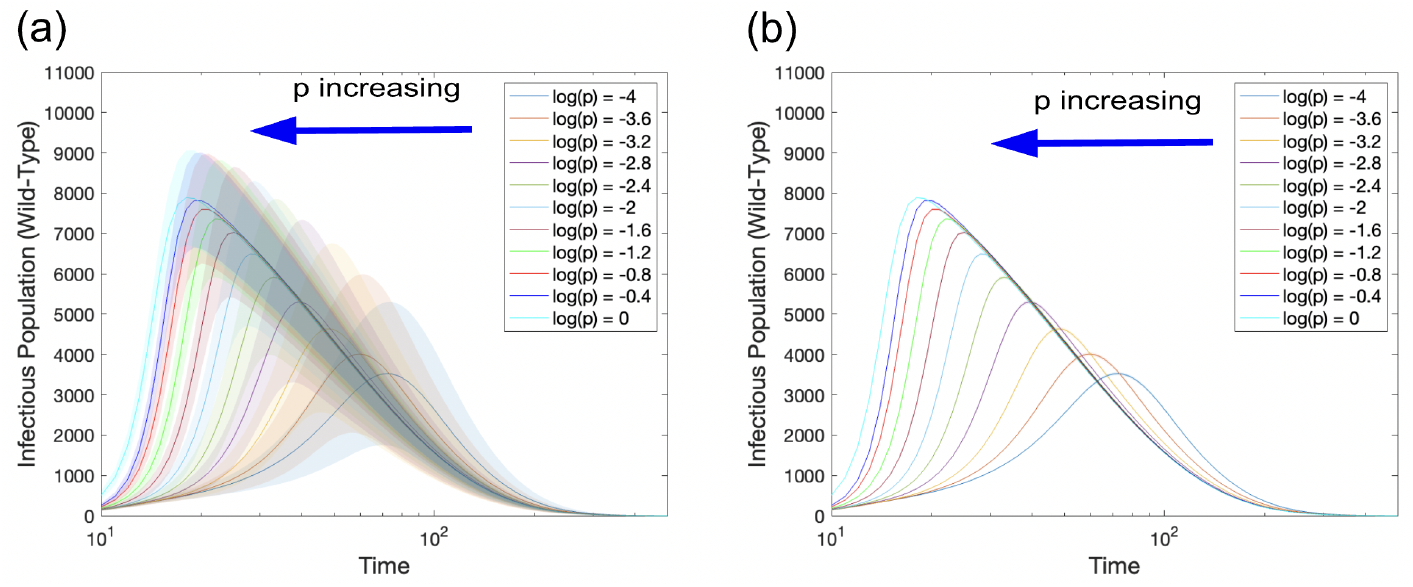
Mean wild-type infectious population for many *p* values. In both panels we show the mean wild-type infectious population over 1,999 simulations. In panel (a) the shaded region represents the standard deviation and in panel (b) the shaded region represents the standard error. Parameters are *N* = 10, 000, ⟨*k*⟩ = 100, *β*_*w*_ = .01, *β*_*m*_ = 1.5*β*_*w*_, *γ* = .02, and *p* is varied according to the legend.

**Figure 16:**
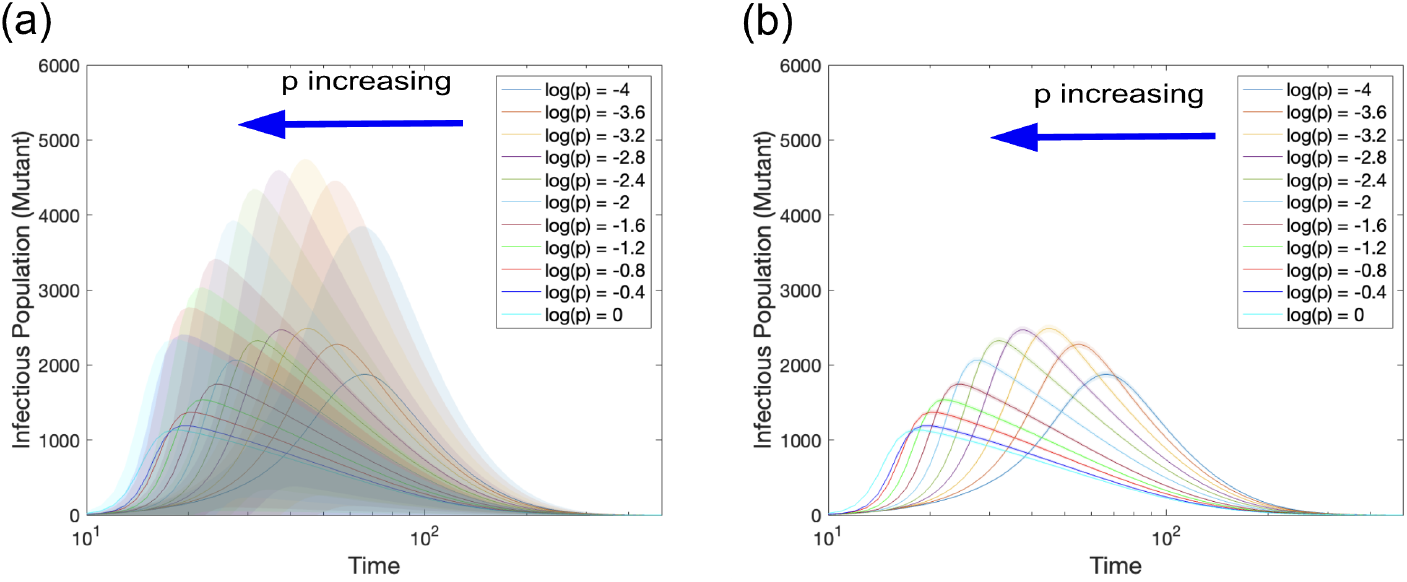
Mean mutant infectious population for many *p* values. In both panels we show the mean mutant infectious population over 1,999 simulations. In panel (a) the shaded region represents the standard deviation and in panel (b) the shaded region represents the standard error. Parameters are *N* = 10, 000, ⟨*k*⟩ = 100, *β*_*w*_ = .01, *β*_*m*_ = 1.5*β*_*w*_, *γ* = .02, and *p* is varied according to the legend.

When *β*_*w*_ increases (while keeping 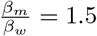, figures 11 and 17) the wild-type reaches a higher maximum at earlier times. For this parameter set the mutants, figures 12 and 18, the mutant peaks also reach a higher maximum at an earlier time step.

**Figure 17:**
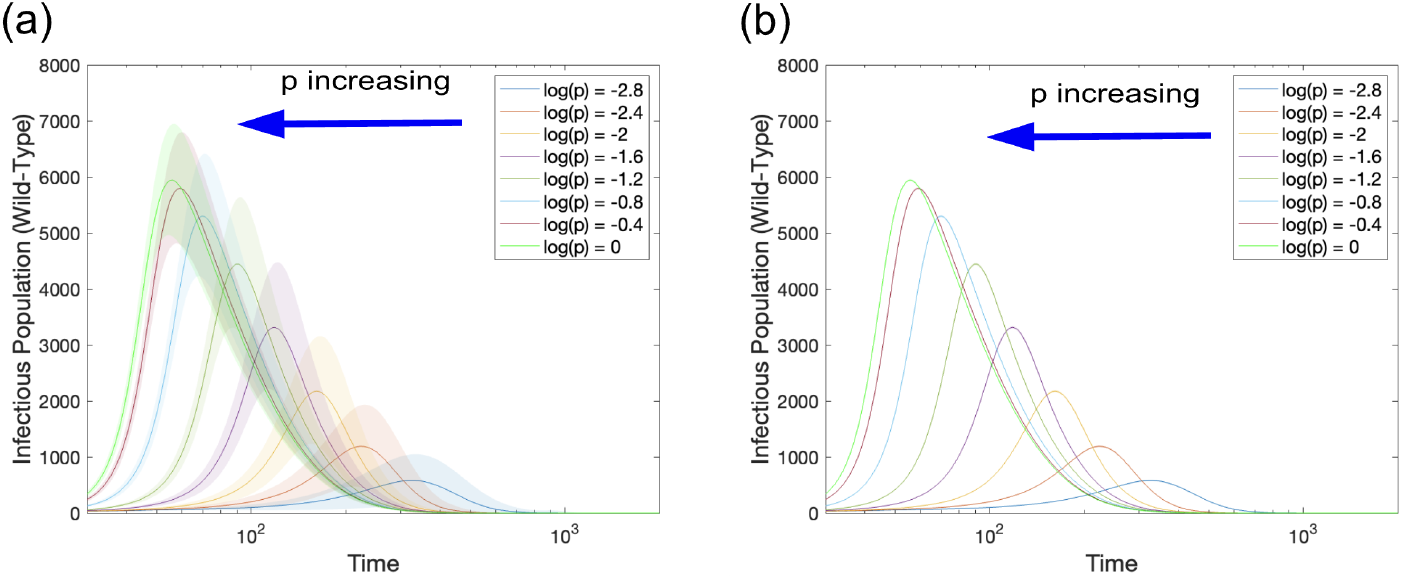
Mean wild-type infectious population for many *p* values. In both panels we show the mean wild-type infectious population over 1,999 simulations. In panel (a) the shaded region represents the standard deviation and in panel (b) the shaded region represents the standard error. Parameters are *N* = 10, 000, ⟨*k*⟩ = 14, *β*_*w*_ = .02, *β*_*m*_ = 1.5*β*_*w*_, *γ* = .02, and *p* is varied according to the legend.

**Figure 18:**
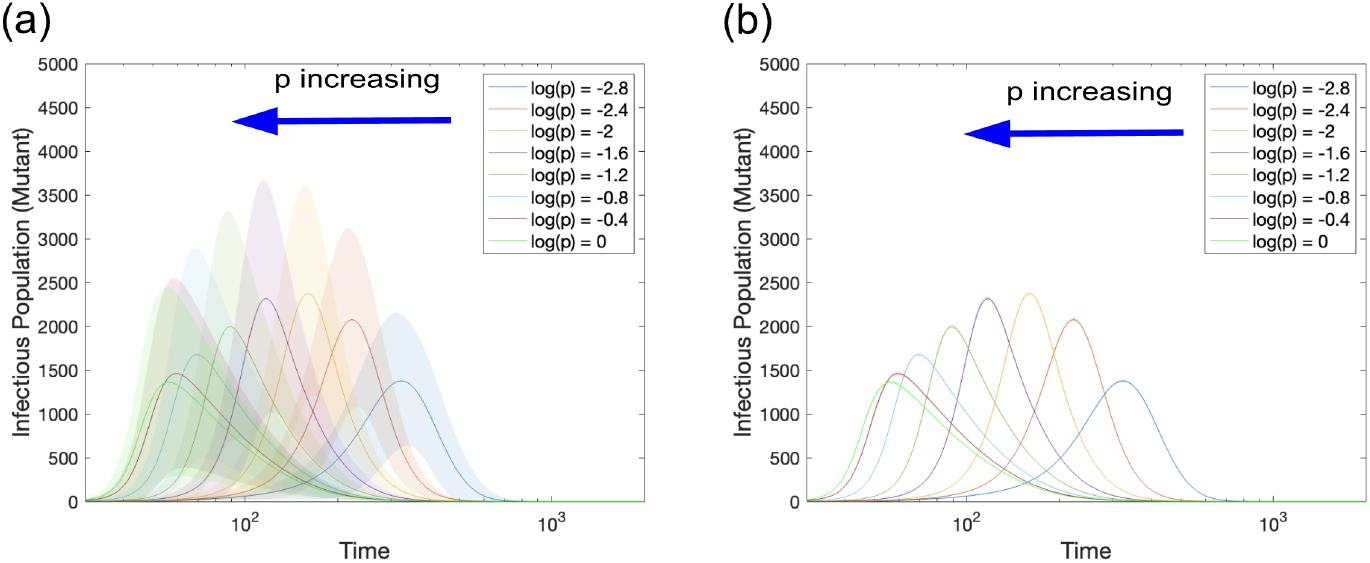
Mean mutant infectious population for many *p* values. In both panels we show the mean mutant infectious population over 1,999 simulations. In panel (a) the shaded region represents the standard deviation and in panel (b) the shaded region represents the standard error. Parameters are *N* = 10, 000, ⟨*k*⟩ = 14, *β*_*w*_ = .02, *β*_*m*_ = 1.5*β*_*w*_, *γ* = .02, and *p* is varied according to the legend.

When the mutant advantage increases to 2 (while keeping *β*_*w*_ = .01, figures 11 and 19), the wild-type achieves lower peaks, while still being monotone increasing as *p* increases. For the mutants, figures 12 and 20, the peaks are larger as we would expect.

**Figure 19:**
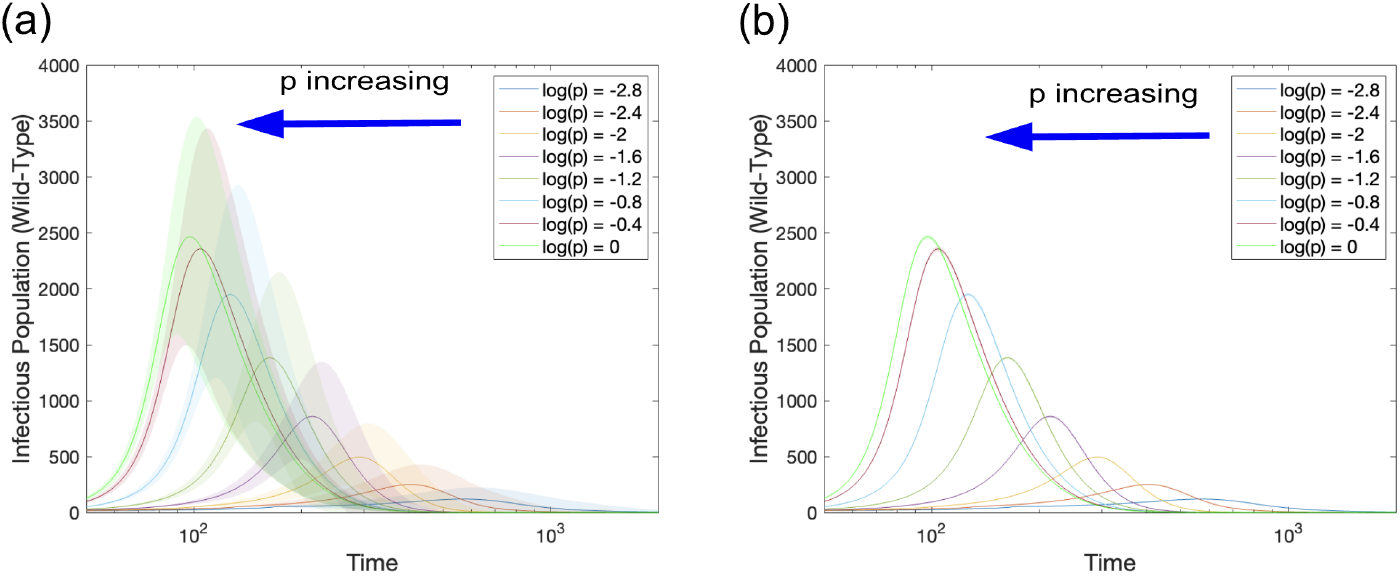
Mean wild-type infectious population for many *p* values. In both panels we show the mean wild-type infectious population over 1,999 simulations. In panel (a) the shaded region represents the standard deviation and in panel (b) the shaded region represents the standard error. Parameters are *N* = 10, 000, ⟨*k*⟩ = 14, *β*_*w*_ = .01, *β*_*m*_ = 2*β*_*w*_, *γ* = .02, and *p* is varied according to the legend.

**Figure 20:**
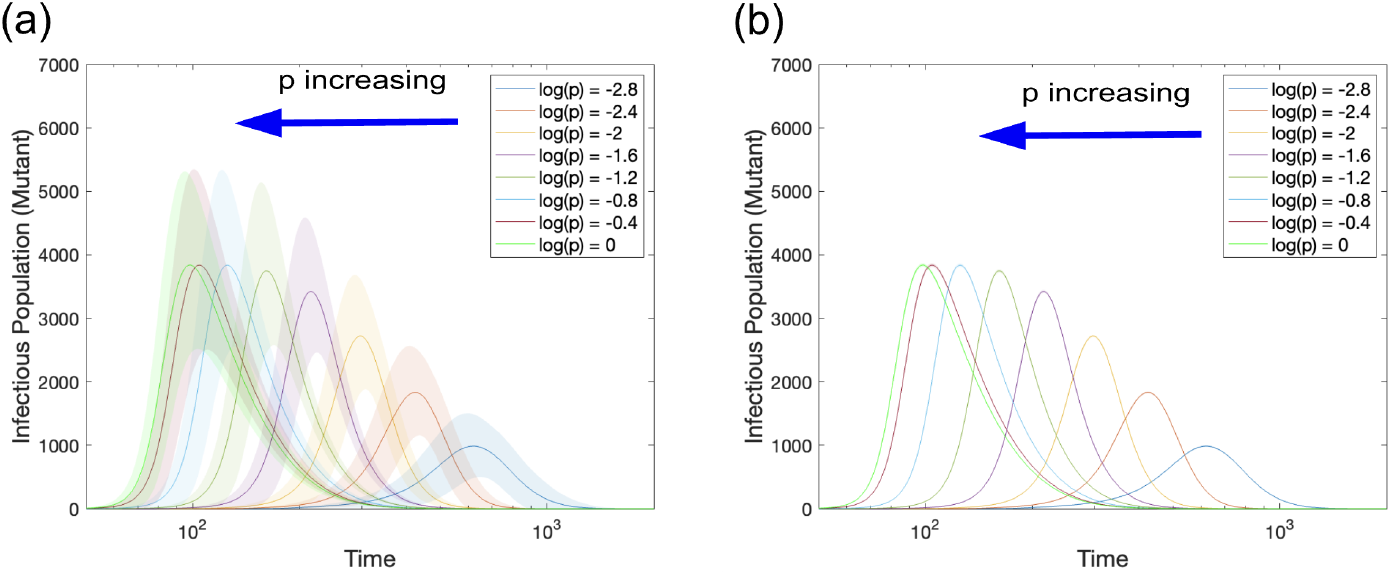
Mean mutant infectious population for many *p* values. In both panels we show the mean mutant infectious population over 1,999 simulations. In panel (a) the shaded region represents the standard deviation and in panel (b) the shaded region represents the standard error. Parameters are *N* = 10, 000, ⟨*k*⟩ = 14, *β*_*w*_ = .01, *β*_*m*_ = 2*β*_*w*_, *γ* = .02, and *p* is varied according to the legend.

When both *β*_*w*_ increases and advantage increases, figures 11 and 21, the peak of the wild-type decreases. Atthe same time, for the mutant, figures 12 and 22, the peak mean infectivity increases.

**Figure 21:**
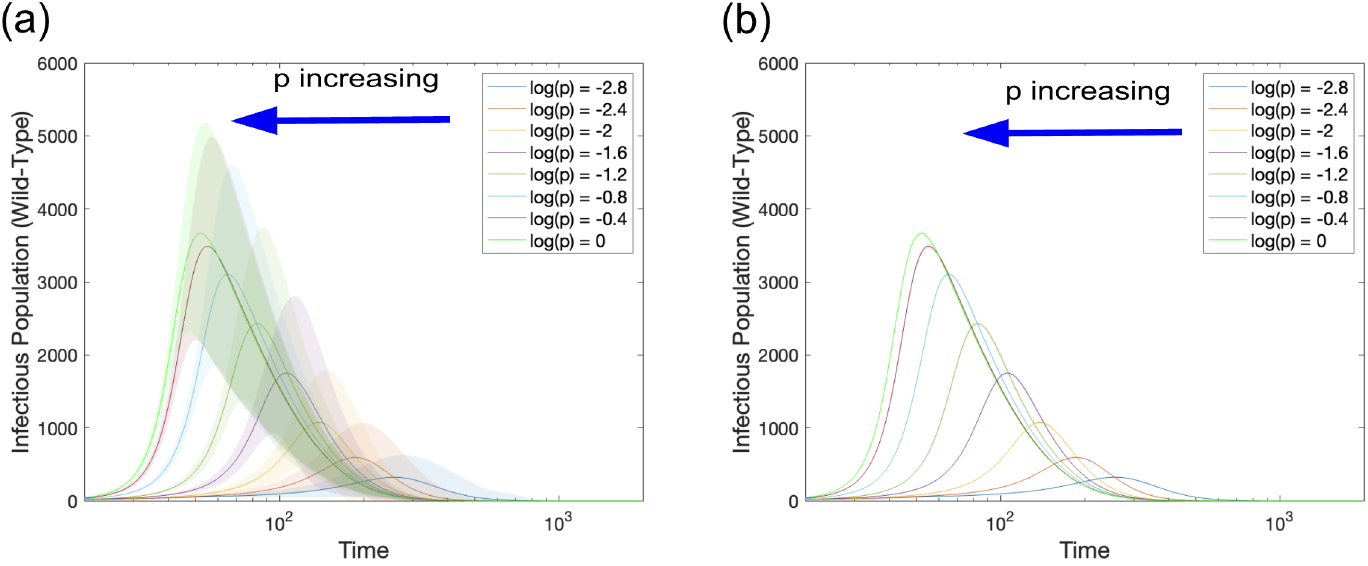
Mean wild-type infectious population for many *p* values. In both panels we show the mean wild-type infectious population over 1,999 simulations. In panel (a) the shaded region represents the standard deviation and in panel (b) the shaded region represents the standard error. Parameters are *N* = 10, 000, ⟨*k*⟩ = 14, *β*_*w*_ = .02, *β*_*m*_ = 2*β*_*w*_, *γ* = .02, and *p* is varied according to the legend.

**Figure 22:**
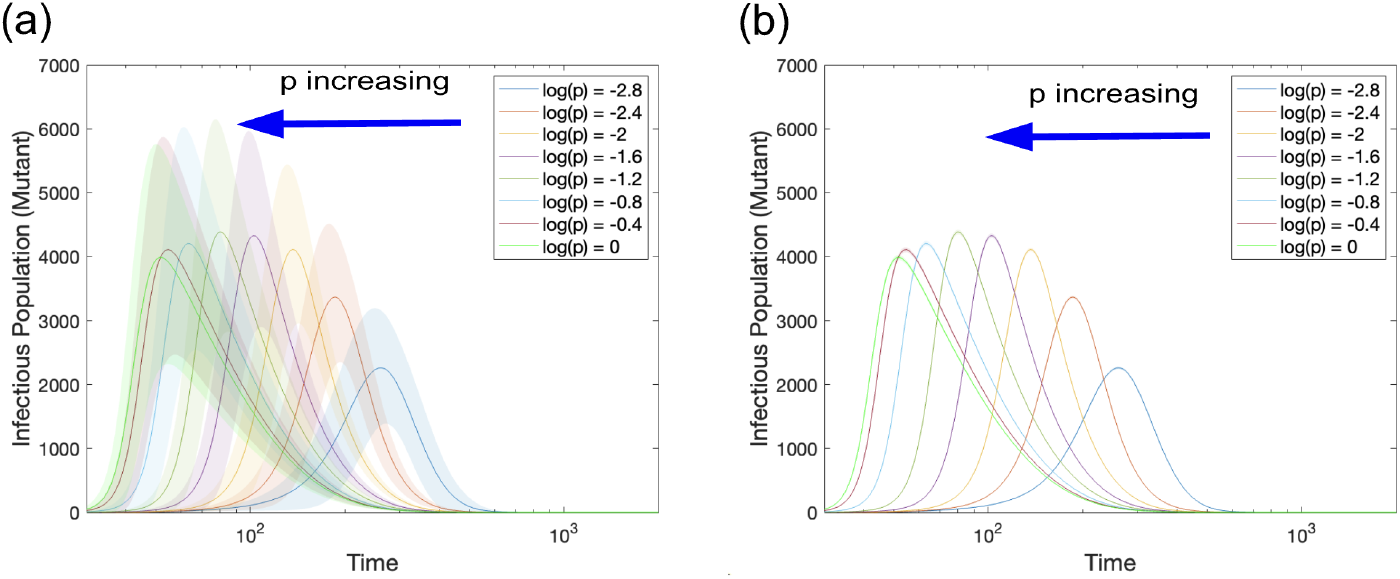
Mean mutant infectious population for many p values. In both panels we show the mean mutant infectious population over 1,999 simulations. In panel (a) the shaded region represents the standard deviation and in panel (b) the shaded region represents the standard error. Parameters are N = 10, 000, ⟨k⟩ = 14, β_w_ = .02, β_m_ = 2β_w_, γ = .02, and p is varied according to the legend.

## Notes

### Competing Interest Statement

The authors have declared no competing interest.

### Funding Statement

This study was partially supported by NSF 2435484, NSF 2438986

## References

[1] COVID-19 Data Repository by the Center for Systems Science and Engineering (CSSE) at Johns Hopkins University. https://github.com/CSSEGISandData/COVID-19. Accessed: 2024-03-30.

[2] Layered SIRS.

[3] HK Alexander and T Day. Risk factors for the evolutionary emergence of pathogens. Journal of The Royal Society Interface, 7(51):1455–1474, 2010.

[4] Christine J Atherstone, Sarah Anne J Guagliardo, Anthony Hawksworth, Kevin O’Laughlin, Kimberly Wong, Michelle L Sloan, Olga Henao, Carol Y Rao, Peter D McElroy, and Sarah D Bennett. SIRS-19 epidemiology during delta variant dominance period in 45 high-income countries, 2020–2021. Emerging Infectious Diseases, 29(9):1757, 2023.

[5] Gábor Bálint, Barbara Vörös-Horváth, and Aleksandar Széchenyi. Omicron: increased transmissibility and decreased pathogenicity. Signal Transduction and Targeted Therapy, 7(1):151, 2022.

[6] Justin Balthrop, Stephanie Forrest, Mark EJ Newman, and Matthew M Williamson. Technological networks and the spread of computer viruses. Science, 304(5670):527–529, 2004.

[7] Patrick Berche. The spanish flu. La Presse Médicale, 51(3):104127, 2022.

[8] Martin Bicher, Claire Rippinger, and Niki Popper. Time dynamics of the spread of virus mutants with increased infectiousness in Austria. Ifac-papersonline, 55(20):445–450, 2022.

[9] Laura Boyle, Sofia Hletko, Jenny Huang, June Lee, Gaurav Pallod, Hwai-Ray Tung, and Richard Durrett. Selective sweeps in SARS-CoV-2 variant competition. Proceedings of the National Academy of Sciences, 119(47):e2213879119, 2022.

[10] Moez Draief, Ayalvadi Ganesh, and Laurent Massoulié. Thresholds for virus spread on networks. In Proceedings of the 1st international conference on Performance evaluation methodolgies and tools, pages 51–es, 2006.

[11] Rebecca Earnest, Rockib Uddin, Nicholas Matluk, Nicholas Renzette, Sarah E Turbett, Katherine J Siddle, Christine Loreth, Gordon Adams, Christopher H Tomkins-Tinch, Mary E Petrone, et al. Comparative transmissibility of SARS-CoV-2 variants delta and alpha in New England, USA. Cell Reports Medicine, 3(4), 2022.

[12] Rashad Eletreby, Yong Zhuang, Kathleen M Carley, Osman Ya?gan, and H Vincent Poor. The effects of evolutionary adaptations on spreading processes in complex networks. Proceedings of the National Academy of Sciences, 117(11):5664–5670, 2020.

[13] Isabel Gordo, M Gabriela M Gomes, Daniel G Reis, and Paulo RA Campos. Genetic diversity in the SIR model of pathogen evolution. PloS one, 4(3):e4876, 2009.

[14] Elena Gubar and Quanyan Zhu. Optimal control of influenza epidemic model with virus mutations. In 2013 European Control Conference (ECC), pages 3125–3130. IEEE, 2013.

[15] Dun Han and Xiao Wang. Vaccination strategies and virulent mutation spread: A game theory study. Chaos, Solitons & Fractals, 176:114106, 2023.

[16] Matthew Hartfield and Samuel Alizon. Within-host stochastic emergence dynamics of immune-escape mutants. PLoS computational biology, 11(3):e1004149, 2015.

[17] Haijun Hu, Xupu Yuan, Lihong Huang, and Chuangxia Huang. Global dynamics of an SIRS model with demographics and transfer from infectious to susceptible on heterogeneous networks. Math. Biosci. Eng, 16(5):5729– 5749, 2019.

[18] Jan Humplik, Alison L Hill, and Martin A Nowak. Evolutionary dynamics of infectious diseases in finite populations. Journal of theoretical biology, 360:149–162, 2014.

[19] Shevin T Jacob, Ian Crozier, William A Fischer, Angela Hewlett, Colleen S Kraft, Marc-Antoine de La Vega, Moses J Soka, Victoria Wahl, Anthony Griffiths, Laura Bollinger, et al. Ebola virus disease. Nature reviews Disease primers, 6(1):13, 2020.

[20] Hildeberto Jardón-Kojakhmetov, Christian Kuehn, Andrea Pugliese, and Mattia Sensi. A geometric analysis of the SIRS epidemiological model on a homogeneous network. Journal of mathematical biology, 83:1–38, 2021.

[21] Jonas S Juul and Steven H Strogatz. Descendant distributions for the impact of mutant contagion on networks. Physical Review Research, 2(3):033005, 2020.

[22] Matthew J Keeling. The effects of local spatial structure on epidemiological invasions. Proceedings of the Royal Society of London. Series B: Biological Sciences, 266(1421):859–867, 1999.

[23] Young Rock Kim, Yong-Jae Choi, and Youngho Min. A model of SIRS-19 pandemic with vaccines and mutant viruses. Plos one, 17(10):e0275851, 2022.

[24] István Z Kiss, Joel C Miller, Péter L Simon, et al. Mathematics of epidemics on networks. Cham: Springer, 598(2017):31, 2017.

[25] Natalia L Komarova, Asma Azizi, and Dominik Wodarz. Network models and the interpretation of prolonged infection plateaus in the COVID19 pandemic. Epidemics, 35:100463, 2021.

[26] Trystan Leng and Matt J Keeling. Improving pairwise approximations for network models with susceptible-infected-susceptible dynamics. Journal of Theoretical Biology, 500:110328, 2020.

[27] Gabriel E Leventhal, Alison L Hill, Martin A Nowak, and Sebastian Bonhoeffer. Evolution and emergence of infectious diseases in theoretical and real-world networks. Nature communications, 6(1):6101, 2015.

[28] Chun-Hsien Li, Chiung-Chiou Tsai, and Suh-Yuh Yang. Analysis of epidemic spreading of an SIRS model in complex heterogeneous networks. Communications in Nonlinear Science and Numerical Simulation, 19(4):1042–1054, 2014.

[29] Jia Li, Yican Zhou, Zhien Ma, and James M Hyman. Epidemiological models for mutating pathogens. SIAM Journal on Applied Mathematics, 65(1):1–23, 2004.

[30] Xue Lin and Qiang Jiao. The equilibrium analysis for competitive spreading over networks with mutations. IEEE Control Systems Letters, 2024.

[31] Ying Liu and Joacim Rocklöv. The reproductive number of the Delta variant of SARS-CoV-2 is far higher compared to the ancestral SARS-CoV-2 virus. Journal of travel medicine, 28(7):taab124, 2021.

[32] Joel C Miller and Istvan Z Kiss. Epidemic spread in networks: Existing methods and current challenges. Mathematical modelling of natural phenomena, 9(2):4–42, 2014.

[33] Hamieh Mohamad, Doumit Mary, Toufaily Joumana, and Hamieh Tayssir. Study of genetic mutations and dynamic spread of SARS-CoV-2 pandemic and prediction of its evolution according to the SIR model. arXiv preprint arXiv:2011.06694, 2020.

[34] Stephen S Morse, Jonna AK Mazet, Mark Woolhouse, Colin R Parrish, Dennis Carroll, William B Karesh, Carlos Zambrana-Torrelio, W Ian Lip-kin, and Peter Daszak. Prediction and prevention of the next pandemic zoonosis. The Lancet, 380(9857):1956–1965, 2012.

[35] Didier Musso and Duane J Gubler. Zika virus. Clinical microbiology reviews, 29(3):487–524, 2016.

[36] Nam P Nguyen, Guanhua Yan, My T Thai, and Stephan Eidenbenz. Containment of misinformation spread in online social networks. In Proceedings of the 4th Annual ACM Web Science Conference, pages 213–222, 2012.

[37] Jonathan M Read and Matt J Keeling. Disease evolution on networks: the role of contact structure. Proceedings of the Royal Society of London. Series B: Biological Sciences, 270(1516):699–708, 2003.

[38] Ganna Rozhnova and Ana Nunes. SIRS dynamics on random networks: Simulations and analytical models. In Complex Sciences: First International Conference, Complex 2009, Shanghai, China, February 23-25, 2009. Revised Papers, Part 1 1, pages 792–797. Springer, 2009.

[39] Sten Rüdiger, Anton Plietzsch, Francesc Sagués, Igor M Sokolov, and Jürgen Kurths. Epidemics with mutating infectivity on small-world networks. Scientific reports, 10(1):5919, 2020.

[40] M Ali Saif. Epidemic threshold for the SIRS model on the networks. Physica A: Statistical Mechanics and its Applications, 535:122251, 2019.

[41] AO Sangotola. A two strain mutation model with temporary and permanent recovery. International Journal of Mathematical Sciences and Optimization: Theory and Applications, 8(1):37–48, 2022.

[42] Reinhard Schlickeiser and Martin Kröger. Determination of a key pandemic parameter of the SIR-epidemic model from past COVID-19 mutant waves and its variation for the validity of the Gaussian evolution. Physics, 5(1):205–214, 2023.

[43] Fabian Jan Schwarzendahl, Jens Grauer, Benno Liebchen, and Hartmut Löwen. Mutation induced infection waves in diseases like COVID-19. Scientific Reports, 12(1):9641, 2022.

[44] Zhi-Gang Shao, Zhi-Jie Tan, Xian-Wu Zou, and Zhun-Zhi Jin. Epidemics with pathogen mutation on small-world networks. Physica A: Statistical Mechanics and Its Applications, 363(2):561–566, 2006.

[45] Masaharu Takahashi, Satoshi Kunita, Tsutomu Nishizawa, Hiroshi Ohnishi, Putu Prathiwi Primadharsini, Shigeo Nagashima, Kazumoto Murata, and Hiroaki Okamoto. Infection dynamics and genomic mutations of hepatitis E virus in naturally infected pigs on a farrow-to-finish farm in Japan: A survey from 2012 to 2021. Viruses, 15(7):1516, 2023.

[46] Kübra Taninmiş, Necati Aras, I. Kuban Altinel, and Evren Güney. Minimizing the misinformation spread in social networks. Iise Transactions, 52(8):850–863, 2020.

[47] Erik Volz. SIR dynamics in random networks with heterogeneous connectivity. Journal of mathematical biology, 56:293–310, 2008.

[48] Xiao Fan Wang and Guanrong Chen. Complex networks: small-world, scale-free and beyond. IEEE Circuits and Systems Magazine, 3(1):6–20, 2003.

[49] Duncan J Watts and Steven H Strogatz. Collective dynamics of ‘small-world’ networks. Nature, 393(6684):440–442, 1998.

[50] Gary Wong, Shihua He, Anders Leung, Wenguang Cao, Yuhai Bi, Zirui Zhang, Wenjun Zhu, Liang Wang, Yuhui Zhao, Keding Cheng, et al. Naturally occurring single mutations in Ebola virus observably impact infectivity. Journal of Virology, 93(1):10–1128, 2019.

[51] Fan Wu, Su Zhao, Bin Yu, Yan-Mei Chen, Wen Wang, Zhi-Gang Song, Yi Hu, Zhao-Wu Tao, Jun-Hua Tian, Yuan-Yuan Pei, et al. A new coronavirus associated with human respiratory disease in China. Nature, 579(7798):265–269, 2020.

[52] Qingchu Wu and Xinchu Fu. Dynamics of competing strains with saturated infectivity and mutation on networks. Journal of Biological Systems, 24(02n03):257–273, 2016.

[53] Chengyi Xia, Li Wang, Shiwen Sun, and Juan Wang. An SIR model with infection delay and propagation vector in complex networks. Nonlinear Dynamics, 69:927–934, 2012.

[54] Ling Yuan, Xing-Yao Huang, Zhong-Yu Liu, Feng Zhang, Xing-Liang Zhu, Jiu-Yang Yu, Xue Ji, Yan-Peng Xu, Guanghui Li, Cui Li, et al. A single mutation in the prM protein of Zika virus contributes to fetal microcephaly. Science, 358(6365):933–936, 2017.

[55] Leyi Zheng and Longkun Tang. A node-based SIRS epidemic model with infective media on complex networks. Complexity, 2019(1):2849196, 2019.

